# Interpretable Speech Features vs. DNN Embeddings: What to Use in the Automatic Assessment of Parkinson’s Disease in Multi-lingual Scenarios

**DOI:** 10.1101/2023.05.29.23290697

**Authors:** Anna Favaro, Yi-Ting Tsai, Ankur Butala, Thomas Thebaud, Jesús Villalba, Najim Dehak, Laureano Moro-Velázquez

**Affiliations:** Department of Electrical and Computer Engineering, The Johns Hopkins University, Baltimore, 21218, MD, US; Department of Neurology, The Johns Hopkins University, Baltimore, 21218, MD, US; Department of Psychiatry and Behavioral Sciences, The Johns Hopkins University, Baltimore, 21218, MD, US

**Keywords:** Parkinson’s disease, Machine learning, Deep learning, Interpretable features, Speech

## Abstract

Individuals with Parkinson’s disease (PD) develop speech impairments that deteriorate their communication capabilities. Speech-based approaches for PD assessment rely on feature extraction for automatic classification or detection. It is desirable for these features to be interpretable to facilitate their development as diagnostic tools in clinical environments. However, many studies propose detection techniques based on non-interpretable embeddings from Deep Neural Networks since these provide high detection accuracy, and do not compare them with the performance of interpretable features for the same task. The goal of this work was twofold: providing a systematic comparison between the predictive capabilities of models based on interpretable and non-interpretable features and exploring the language robustness of the features themselves. As interpretable features, prosodic, linguistic, and cognitive descriptors were employed. As non-interpretable features, x-vectors, Wav2Vec 2.0, HuBERT, and TRILLsson representations were used. To the best of our knowledge, this is the first study applying TRILLsson and HuBERT to PD detection. Mono-lingual, multi-lingual, and cross-lingual machine learning experiments were conducted on six data sets. These contain speech recordings from different languages: American English, Castilian Spanish, Colombian Spanish, Italian, German, and Czech. For interpretable feature-based models, the mean of the best F1-scores obtained from each language was 81% in mono-lingual, 81% in multi-lingual, and 71% in cross-lingual experiments. For non-interpretable feature-based models, instead, they were 85% in mono-lingual, 88% in multi-lingual, and 79% in cross-lingual experiments. On one hand, models based on non-interpretable features outperformed interpretable ones, especially in cross-lingual experiments. Among the non-interpretable features used, TRILLsson provided the most stable and accurate results across tasks and data sets. Conversely, the two types of features adopted showed some level of language robustness in multi-lingual and cross-lingual experiments. Overall, these results suggest that interpretable feature-based models can be used by clinicians to evaluate the evolution and the possible deterioration of the speech of patients with PD, while non-interpretable feature-based models can be leveraged to achieve higher detection accuracy.

**Highlights:**

- Both interpretable and non-interpretable features displayed robust behaviors.
- Models based on non-interpretable features outperformed interpretable ones.
- Interpretable feature-based models provide insights into speech and language deterioration.
- Non-interpretable feature-based models can be used to achieve higher detection accuracy.

## 1. Introduction

Parkinson’s disease (PD) is the second most common neurodegenerative disease after Alzheimer’s. It results from the gradual neuronal death in the substantia nigra [1], closely related to the production of dopamine neurotransmitters. Clinical diagnosis is based on cardinal motor signs [2] and other non-motor indicators (physiological and cognitive manifestations). The average time to diagnose PD in clinical setting can reach 2.9 years, with diagnostic accuracy around 80.6% (95% Credible Interval [CrI] 75.2%-85.3%) [3, 4]. In addition, tracking PD progression includes the adoption of subjective rating scales (e.g., Hoehn & Yahr [5]) that are considered to have low sensitivity and inter-rater reliability at the mild end of the symptom severity spectrum [6, 7]. This is due, in part, to the variable and subtle nature of the early symptomatic presentation. In this respect, novel technologies can speed up the diagnosis process, help test treatments before largely irreversible brain damage, and slow down the disorder’s progression. Voice disorders affect approximately 70 to 90% of individuals with PD [8, 9, 10, 11]. Initial studies designing interpretable features for PD evaluation have focused on representing phonatory, articulatory, and prosodic aspects such as perturbations of fundamental frequency (jitter), perturbations of amplitude (shimmer), voice onset time, vowel space area, number of pauses, standard deviation of pitch and intensity, and formats [12, 13, 14, 15, 16, 17]. Others used linguistic descriptors to represent word dysfluencies, vocabulary richness, and word repetitions [18, 19, 20]. These features were usually adopted to classify the speech of subjects in PD and healthy controls (HC) with different classification techniques (e.g., Support Vector Machines, k-nearest neighbors, multilayer perceptrons) [21, 22, 23, 24]. Even though designing interpretable features is desirable in clinical practice, this process can entail significant effort in terms of computation and domain expertise. Domain expertise is required to provide the definition of interpretability for the domain of application and to design the features for machine learning (ML) [25]. Over the last few years, Deep Neural Networks (DNNs) have been extensively adopted in PD detection. In particular, Convolutional Neural Networks (CNNs) have been used to extract features from spectrograms or phonological features from Mel-Frequency Cepstral Coefficients (MFCCs) [26, 27, 28]. Other studies have explored the use of speaker recognition technologies such as i-vectors and x-vectors in parameterizing articulatory, prosodic, and phonatory information characteristic of speakers with PD [29, 30, 31, 32]. The use of Deep Learning (DL) is increasing in neurology, pathology, and medicine in general [33, 34, 35]. However, the lack of transparency and accountability of predictive models can have severe consequences and generally poor use of limited valuable resources in medicine and other domains [36].

Among the existing studies developing speech-based features for PD detection, few of them extended their analysis to a multi-lingual cohort. Hazan et al. [37] focused on the automatic detection of PD in English and German. They reached 85% accuracy using Support Vector Machines (SVM) in monolingual and 75% in multi-lingual experiments, respectively. Orozco-Arroyave et al. [38] performed cross-lingual experiments with Spanish, German, and Czech data sets. Accuracy ranges from 60% to 99%, depending on the language combination and the ratio of training/testing data. Regarding a single-language evaluation, the accuracies were between 85% and 99%. The most valuable features were based on MFCCs and Bark band energies calculated from unvoiced segments of reading text. Moro-Veĺazquez et al. [39] reached accuracies between 75% and 82% when classifying PD individuals in cross-lingual experiments considering Castilian Spanish, Colombian Spanish, and Czech data sets. For this purpose, they used an approach based on phonemic grouping. Plosives, vowels, and fricatives were the most relevant acoustic segments for the detection of PD. Using text-dependent utterances as a speech task led to the highest performance. Vásquez Correa et al. [40, 27] used CNNs and a transfer learning strategy to classify PD in Spanish, German, and Czech. Accuracies varied from 70% to 77%. Classifying German- and Czech-speaking participants with PD was more accurate when pre-training the model on the Spanish data set. Kovac et al. [41] showed that particular language tasks used for PD detection (e.g., monologue) are very language-dependent and that just a few features (e.g., prosodic) have significant discrimination power across the Czech and US-English language. The classification accuracy reported in their experiments dropped from 72–73% to 67% when moving from single-language modeling to multi-lingual one. Differently, Kovac et al. [42] analyzed acoustic features of Czech, American, Israeli, Colombian, and Italian PD and HC participants using correlation and statistical tests. Classification accuracies ranged from 67% to 85% in mono-lingual experiments and from 53% to 79% in cross-lingual experiments using leave-one-language-out. Performance varied depending on the language.

Most of the mentioned studies have different limitations. Some only analyzed acoustic or linguistic descriptors, which limits the characterization of PD to a unique domain. Others only leveraged DL techniques to detect PD from speech, which is not entirely desirable in clinical applications, given their limited interpretability. Others only considered a single language task (e.g., sustained vowel phonation, diadochokinetic task) or only one or two languages simultaneously. Thus, the task of PD detection from language and speech requires adopting a broader set of languages, focusing on a more comprehensive set of descriptors, and collecting speech samples from different tasks to deliver an exhaustive characterization of the disorder.

The main goal of this work is to perform a systematic comparison between interpretable feature-based models (IFMs) and non-interpretable feature-based models (NIFMs) for the detection of PD from speech. As previously discussed by Rudin [25], there is a wide belief that using more complex models leads to greater accuracy in predictions, which suggests that a complex *black box* is necessary for achieving optimal results. However, this notion is frequently disproven, especially in cases where the data is structured and has natural, meaningful features to be effectively represented. As shown by some works in data science [43, 44, 45], in such situations, the performance of more complex classifiers like deep neural networks tend to be comparable to that of simpler classifiers such as logistic regression and decision lists after preprocessing. In our study, we wanted to assess whether simpler models based on meaningful and interpretable features can achieve a performance comparable to more complex models based on novel DNN-based representations such as TRILLsson [46], Wav2Vec 2.0 [47], and HuBERT [48]. Moreover, we intended to evaluate which of the two typologies of representations encodes more suitable information for the task of PD detection and to which extent the representations adopted are language robust. To do so, we performed mono-lingual, multi-lingual, and cross-lingual experiments using three different speech tasks and six corpora simultaneously. To the best of our knowledge, there are no studies that conducted a comparison between the predictive capabilities of interpretable and non-interpretable speech-derived features in PD detection across languages.

## 2. Materials

Mono-lingual, multi-lingual, and cross-lingual experiments comprised six corpora: American English, Castilian Spanish, Colombian Spanish, German, Italian, and Czech. The data sets used for each language are described below. From each data set, we employed speech recordings from three types of tasks (when available): spontaneous speech (SS) (e.g., monologue, picture description), reading passage (RP), and text-dependent utterances (TDUs) (short read sentences). Table 1 summarizes the typology of tasks adopted in this study and the data sets in which these tasks are available. Demographic and disease severity statistics of American English, German and Italian corpora are reported in Table 2, while those of Castillian, Colombian, and Czech are reported in Table 3.

**Table 1:** Typologies of tasks analyzed with the corresponding number (#) of samples available in each data set. Abbreviations: SS, Spontaneous Speech; RP, Read Passage; TDU, Text Dependent Utterance.

| Data set | Spontaneous speech | Read passage | Text dependent utterance |
| --- | --- | --- | --- |
| NLS | 1 | 2 | 0 |
| Neurovoz | 1 | 0 | 6 |
| GermanPD | 1 | 1 | 5 |
| CzechPD | 1 | 1 | 0 |
| GITA | 1 | 1 | 6 |
| ItalianPVS | 0 | 2 | 10 |

**Table 2:** Demographic and disease severity statistics of NLS, GermanPD, and ItalianPVS data sets. Ages are expressed in years. *NLS contains global values of Movement Disorder Society UPDRS part III (i.e., movement examination); GermanPD contains global values of UPDRS part III (i.e., movement examination). The ItalianPVS corpus only contains UPDRS-III.I (i. e. speech examination). The Hoehn and Yahr* scale is available only for NLS, and GermanPD, but not for ItalianPVS.

| Data set | NLS |  |  |  | German PD |  |  |  | ItalianPVS |  |  |  |
| --- | --- | --- | --- | --- | --- | --- | --- | --- | --- | --- | --- | --- |
| Gender | Female |  | Male |  | Female |  | Male |  | Female |  | Male |  |
| Group | PD | HC | PD | HC | PD | HC | PD | HC | PD | HC | PD | HC |
| Number of Subjects | 9 | 16 | 14 | 11 | 41 | 44 | 47 | 44 | 9 | 12 | 19 | 10 |
| Age, Average | 68.00 | 63.31 | 66.62 | 66.09 | 67.23<br>(9.7) | 62.6<br>(15.2) | 66.7<br>(8.7) | 63.8<br>(12.7) | 64.3<br>(12.2) | 65.3<br>(4.1) | 68.6<br>(6.4) | 69.3<br>(5.6) |
| Age Range | 50-75 | 26-83 | 55-79 | 41-79 | 27-84 | 28-85 | 44-82 | 26-83 | 40-80 | 60-72 | 50-77 | 60-77 |
| UPDRS-III*, Average (SD) | 25.33<br>(9.72) | – | 27.71<br>(13.63) | – | 23.3<br>(12.0) | – | 22.1<br>(9.9) | – | – | – | – | – |
| UPDRS-III.I Average (SD) | 0.88<br>(0.56) | – | 0.83<br>(0.68) | – | – | – | – | – | 1.10<br>(1.3) | – | 1.00<br>(1.00) | – |
| Hoehn and Yahr* Scale Average (SD) | 2.37<br>(0.41) | – | 2.23<br>(0.37) | – | 2.61<br>(0.83) | – | 2.59<br>(0.60) | – | – | – | – | – |
| Years Since Diagnosis (SD) | – | – | – | – | 6.47<br>(5.83) | – | 6.59<br>(4.93) | – | – | – | – | – |

**Table 3:** Demographic and disease severity statistics of Neurovoz, GITA and CzechPD data sets. Ages are expressed in years. *The Neurovoz corpus only contains UPDRS-III, i.e. motor examination; GITA contains global values of Movement Disorder Society UPDRS; CzechPD contains global values of UPDRS.

| Data set | Neurovoz |  |  |  | GITA |  |  |  | CzechPD |  |
| --- | --- | --- | --- | --- | --- | --- | --- | --- | --- | --- |
| Gender | Female |  | Male |  | Female |  | Male |  | Male |  |
| Group | PD | Ctrl | PD | Ctrl | PD | Ctrl | PD | Ctrl | PD | Ctrl |
| Number of Subjects | 18 | 18 | 29 | 14 | 25 | 25 | 25 | 25 | 20 | 16 |
| Age, Average (SD) | 70.9 (8.4) | 68.4 (6.0) | 71.9 (12.3) | 66.6 (6.4) | 60.7 (7.3) | 61.4 (7.0) | 61.5 (11.6) | 60.5 (11.6) | 61 (11.7) | 61.8 (12.9) |
| Age Range | 59-86 | 58-83 | 41-88 | 55-77 | 49-75 | 49-76 | 33-81 | 31-86 | 34-83 | 36-80 |
| UPDRS-III*, Average (SD) | 16.9 (11.5) | – | 19.6 (11.8) | – | 37.5 (14.0) | – | 37.7 (22.0) | – | 17.9 (7.1) | – |
| Hoehn and Yahr Scale Average (SD) | 2.30 (0.68) | – | 2.30 (0.80) | – | 2.28 (0.53) | – | 2.34 (0.53) | – | 2.17 (0.45) | – |
| Years Since Diagnosis (SD) | 6.4 (6.4) | – | 7.6 (4.7) | – | 12.6 (11.5) | – | 8.9 (5.9) | – | 2.4 (1.6) | – |

### 2.1. American English

The authors of this study collected a data set called NeuroLogical Signals (NLS) at Johns Hopkins Medicine (JHM). This data set consists of spoken responses from participants who are native speakers of American English. The participants were categorized as either having a neurological disorder or being HC and received treatment and diagnosis from expert neurologists at JHM. All participants underwent informed consent, and the data collection was approved by the Johns Hopkins Medical Institutional Review Board. Due to the COVID-19 pandemic, all participants wore the same type of surgical mask during recordings, which minimally interfered with their jaw and lip movement. Participants with PD continued their usual pharmacological treatment and took dopaminergic medication before the recording session. Speech signals were recorded in acoustically and visually controlled conditions with a computer and microphone. The study included 23 participants with PD and an HC group of 27 participants matched in age with the PD group. None of the participants in the HC group has a history of symptoms related to PD or any other NDs. The subset of tasks analyzed from this corpus consists of a SS task and two RP tasks. The SS task is represented by a Cookie Theft Picture description task [49], in which participants are required to describe the Cookie Theft Picture from the Boston Diagnostic Aphasia Examination [50]. The protocol sets a limit of 60 s on the execution of this task. In the two RPs instead, participants are instructed to read two short passages consisting of 87 and 174 words, respectively, from a printed sheet.

### 2.2. Castilian Spanish

The Neurovoz data set contains recordings from 47 HCs and 32 participants with PD who are native speakers of Castilian Spanish. The Bioengineering and Optoelectronics Group at Universidad Politecnica de Madrid and the Otorhinolaryngology and Neurology Departments of the Gregorio Marañෳon hospital (Madrid, Spain) collected the speech samples. The experimental protocols and methods were approved by the Ethics Committee of Hospital General Universitario Gregorio Marañón in accordance with the Helsinki Declaration [51], and all participants gave informed consent. Before the recording session, a neurologist assessed the neurological state of participants with PD, while a survey was administered to evaluate the neurological condition of HC participants. PD participants were under pharmacological treatment and took dopaminergic medication before the recording session. Speech signals were collected in a room with controlled acoustic characteristics using a headset microphone. Recordings were sampled at 44.1 kHz with a resolution of 16 bits. The data set included recordings from six TDUs and running speech from the description of a picture (SS task). The SS task did not have a time limit. For the TDUs, participants did not read the sentences from a text document but instead listened to them from a recording of a standard speaker and repeated them out loud to reduce noise, reading mistakes, and cognitive load.

### 2.3. Colombian Spanish

The GITA data set was collected by Universidad de Antioquia in Medelĺın, Colombia, and includes speech samples from 50 individuals with PD and 50 age-and sex-matched HC who are native speakers of Colombian Spanish. The data collection adhered to the Helsinki Declaration and was approved by the Ethics Committee of Cĺınica Noel in Medelĺın, and all participants provided informed consent. PD patients were diagnosed by neurology experts and took dopaminergic medication within 3 hours before the start of the recording session. HC participants reported no symptoms related to PD or other NDs. The recordings were conducted in a quiet room using an external microphone at a sampling rate of 44.1 kHz with a resolution of 16 bits. The data set includes three types of speech tasks: a SS task (monologue) where participants describe their typical daily activities, six TDUs, and a phonetically balanced RP. The SS task had no time limit, and the recorded samples were approximately 44 s long.

### 2.4. German

The GermanPD data set comprises speech recordings from 88 participants with PD and 88 HC participants who are native German speakers. The data were collected at the Hospital of Bochum in Germany and approved by the ethics committee of the Ruhr-University Bochum. All participants provided informed consent. The participants with PD received dopaminergic medication before the recording session. Speech samples were collected using commercial audio software and a headset microphone in a quiet room. Recordings were sampled at 16 kHz with a resolution of 16 bits. The subset of tasks analyzed in this study includes ten TDUs, one RP composed of eighty-one words, and a SS task. The protocol did not impose a time limit on the execution of the SS task, and the SS samples last about 33 s. The neurological state of participants with PD was assessed by a neurologist, and none of the HC participants reported symptoms associated with PD or other neurological disorders.

### 2.5. Italian

The ItalianPVS corpus^1^, comprises recordings from 22 elderly HCs, 28 young HCs, and 28 participants diagnosed with PD. All information regarding participant consent is available in their reference papers or dissemination platforms, which are cited in this paper. None of the participants with PD reported any speech or language disorders unrelated to their PD symptoms prior to this study. All participants with PD received dopaminergic medication at the time of the recording. Speech samples were collected in a quiet, echo-free room, using ab external microphone. Recordings were sampled at 16 kHz. The set of tasks analyzed in this study consists of recordings from ten phonetically balanced TDUs and one phonetically balanced RP. Prior to starting the reading tasks, a specialist introduced the participants to the task to be performed, and both the short sentences and the RP were read from a printed sheet.

### 2.6. Czech

The CzechPD data set was collected in the General University Hospital in Prague, Czech Republic. It comprises speech recordings from 20 untreated male participants newly diagnosed with PD and 15 age-matched HC individuals who speak Czech natively. The data collection process adhered to the Helsinki Declaration, and the Ethics Committee of the hospital approved it. Every participant provided informed consent. Participants with PD did not take any antiparkinsonian treatment before recording. The recording session took place in a quiet room. Speech samples were collected with an external condenser microphone and originally sampled at 48 kHz with a resolution of 16 bits. Participants were familiarized with the task and procedure before each recording session. In this study, participants performed various speaking tasks, including a phonetically balanced reading passage and a monologue task, where they spoke about their daily activities, interests, jobs, or family. The SS samples lasted approximately 90 s, and there were no time limits imposed on the recording.

## 3. Methods

Our pipeline employed a standard “feature extraction-classification” scheme. Nested Cross-Validation (NCV) was applied in the experiments to evaluate the two different representation techniques.

### 3.1. Data Pre-processing

All recordings were resampled at 16 kHz as required by the algorithms employed for the feature extraction (see Section 3.2). The resampling was performed using SoX.^2^ We also applied EBU R128 loudness normalization procedure using the Python library *ffmpeg-normalize*.^3^ This type of normalization leads to a more uniform loudness level compared to simple peak-based normalization. Normalized audios were used to extract intensity-related features (see Section 3.2.1). Moreover, whenever a given participant had more than one speech sample within a given task, we concatenated all the recordings to form a single one. In doing this, we trimmed silences at the end and beginning of each recording. We supervised all the recordings in the NLS data set to ensure they had appropriate quality. Criteria for appropriate quality encompassed no distortion, minimal background noise, and a task-related response. When the recordings contained speech from the investigator at the beginning and end, we trimmed the recordings. All the recordings from the SS task were automatically transcribed using OpenAI’s Whisper^4^ [52], an Automatic Speech Recognition (ASR) system trained on 680.000 hours of multi-lingual and multitask supervised data collected from the web. American English transcriptions underwent manual supervision.

### 3.2. Feature Extraction

We adopted two distinct characterizations to represent speech traits connected to the onset and progression of PD. On the one hand, we configured a set of interpretable features, some of them already proposed in our previous works [53, 54, 55], encoding prosodic, linguistic, and cognitive information. A more detailed list of the interpretable features extracted is provided in the Supplementary Material. On the other hand, non-interpretable speech representations, namely neural network embeddings extracted using pre-trained DNNs, were utilized. A summary of the features extracted for both approaches is reported in Table 4.

**Table 4:** Summary of the interpretable and non-interpretable features extracted.

| Feature type | # of features | Feature description |
| --- | --- | --- |
| Interpretable features |  |  |
| Acoustic | 30 | Features based on F0 (e.g., F0-standard deviation) |
|  | 48 | Features based on energy (e.g., energy-contour on voiced segments) |
|  | 24 | Features based on duration (e.g., voiced rate) |
|  | 20 | Feature based on pauses (e.g., mean pause length) |
| Linguistic | 10 | Features based on the part of speech (POS) (e.g., number of verbs) |
|  | 2 | Features based on syntactic phrases (e.g., number of noun phrases) |
| Cognitive | 2 | Feature based on rhythm (i.e., speech rhythm standard deviation) |
|  | 1 | Feature based on informational content (e.g., number of correct informational units) |
| Non interpretable features |  |  |
| X-vectors | – | X-vector embeddings pre-trained on Voxceleb1 and Voxceleb2 |
| TRILLsson | – | TRILLsson1 embeddings distilled on AudioSet and Libri-light from a CAP12 teacher model pre-trained on YT-U |
| Wav2Vec 2.0 | – | Wav2Vec 2.0 embeddings pre-trained on LibriSpeech |
| HuBERT | – | HuBERT embeddings pre-trained on LibriSpeech |

#### 3.2.1. Interpretable Features

##### Prosodic Features

Prosody is one of the most deteriorated speech dimensions in hypokinetic dysarthria, a motor speech disorder associated with PD [56, 57]. The speech of individuals with PD is usually characterized by monopitch, monoloudness, reduced overall loudness, short rushes of speech, speech rate abnormalities, excessive and longer speech pauses, and reduced stress [56, 58, 59, 60]. To extract prosody features based on duration, fundamental frequency, and energy, we used Disvoice^5^, a Python library designed to extract phonological, prosodic, articulatory, and glottal features from speech. To compute loudness variation, we used Parselmouth^6^, a Python library for the Praat software^7^. We also used the Python library DigiPsychProsody^8^, to compute features related to pauses such as total speech time, total pause time, percentage pause time, mean pause duration, silence to speech ratio, and pause variability. This library uses the WebRTC Voice Activity Detector^9^. A total of 125 prosodic features were extracted.

##### Linguistic Features

In some studies, individuals with PD report selective impairments in syntax and semantics, especially in action-verbs and action semantics, with relative preservation of noun processing [61]. In others, individuals with PD exhibit difficulties in both actions and object naming [62] and improved action naming more than object naming upon stimulation of the subthalamic nucleus [63]. To model the syntactic abilities of participants with PD during a SS task, we calculated the frequency of occurrence of different parts of speech (POS), such as nouns, verbs, adjectives, adverbs, numerals, and auxiliaries during spontaneous production. We also measured the syntactic complexity in terms of the number of words, average word length in characters, number of sentences, average sentence length in words, number of noun phrases (NPs), number of verb phrases (VPs), and number of prepositional phrases (PPs). To extract these features, we used the pretrained pipeline for English, German, and Spanish available on Spacy^10^. Since there is no available pipeline for Czech to perform POS tagging and syntactic analysis, we did not perform the linguistic feature extraction on the CzechPD, even though this data set contains a SS task. A total of thirteen linguistic features was extracted.

##### Cognitive Features

Previous works demonstrated that individuals with PD have difficulties with regulating the rhythm and timing of speech during spontaneous production [64, 65]. In our analysis, we modeled the regularity of speech rhythm in terms of the occurrence of the individual words in time. Namely, we measured the time between the starting points of consequent words, and we computed the standard deviation, kurtosis, and skewness of these measurements for each recording. To derive the starting point of each word, we computed word alignment using a modified method of Whisper’s model.^11^ In addition, it has been shown that subjects with PD report a lower informational content than HCs when required to mention the main characters or events represented in a picture [54, 66, 67, 68].

We also represented the informativeness of the narratives in terms of the number of correct informational units (IU). According to [69], an IU is a *word that is intelligible in context, accurate in relation to the picture or topic, and relevant to and informative about the content of the picture(s) or the topic*. In our analysis, we employed the speech transcripts of the recordings collected during the picture description tasks from both NLS and Neurovoz. We considered IUs, the salient events displayed in the picture presented as a stimulus to elicit the narrative [55]. To identify the salient IUs for the CTP task, we leveraged the published checklists for the CTP [70, 71]. Thus, IUs were represented by verbs like *washing*, *drying*, *stealing*, *overflowing*, *trying to help*, *falling*, *wobbling*, *hanging*, *ignoring*, *reaching up*, *asking for cookie*, *laughing*, *standing*. On the other hand, we adopted the verbs *barrer*, *lavar*, *pesar*, *ducharse* to represent the four main actions displayed in the picture used in Neurovoz. A total of four cognitive features were extracted.

#### 3.2.2. Non-interpretable Features

##### X-vectors

These are embedding features extracted from DNNs used for speaker recognition tasks [72]. They are very robust speaker representations, and their network scheme can be adapted to apply to language and emotion recognition as well [73, 74]. Previous studies showed that x-vectors are effective in differentiating between the speech of PD and HC, and they work equally well for early PD detection in bi-class scenarios [31, 32]. More importantly, x-vectors display robustness to language dependency. In the two studies mentioned above, x-vector models were trained on Voxceleb1 [75] to extract speaker embeddings. An excellent performance was achieved when evaluating both French and Spanish PD data sets. Besides the benefit of capturing multi-lingual PD characteristics, x-vectors have large pre-trained models. When it comes to DNN schemes for PD detection, more training data is needed. However, since pre-trained x-vector models can be directly applied without further training, they can be extracted from all the different data sets regardless of the amount of available data. In our analysis, the pre-trained x-vector model^12^ available from SpeechBrain [76] was used. SpeechBrain is an open-source and all-in-one conversational AI toolkit based on PyTorch. This particular x-vector model was trained on both Voxceleb1 (English only) [75], and Voxceleb2 (multi-lingual) [77]. Given a speech recording as input, the model extracts an x-vector of size 512. Recordings need to be sampled at 16 kHz.

##### Self-Supervised Representations

Depending on the domain of application, labeled data are challenging to obtain. Thus, research into pre-training models with unlabelled data has significantly advanced. In this respect, self-supervised learning (SSL) involves training a model on a large amount of unlabelled data and learning an acoustic representation to be used on a downstream task. Most SSL models rely on unsupervised pre-training followed by supervised fine-tuning on a downstream task. The unsupervised pre-training allows a model to benefit from the large amounts of unlabeled data which are readily available. The following approaches were used in this study.

- *TRILLsson.* Paralinguistic representations encode non-lexical elements of communication of speech, such as emotion and tone. In [78], the CAP12 model was introduced. The model extracted state-of-the-art paralinguistic representations from large-scale, fully self-supervised training of a 600*M* + parameter Conformer-based architecture. It has been shown that simple linear classifiers trained on top of time-averaged CAP12 representations outperformed nearly all previous state-of-the-art representations on paralinguistic tasks such as speech emotion recognition, synthetic speech detection, and dysarthria classification. The publicly available versions of CAP12 are TRILLsson models. They use knowledge distillation to derive smaller, faster, and more mobile-friendly architectures than the original CAP12 model [46]. Distillation was performed on the public data sets Audioset [79], and Libri-light [80], with CAP12 as teacher model trained on YT-U [81]. The distilled TRILLsson1 model^13^ was used in this study. Despite being much smaller, TRILLsson models outperformed publicly available representations like TRILL [82] and Wave2Vec2.0 [47] on multiple paralinguistic tasks. Similar to the x-vector model, given a speech recording of any length as input, the TRILLsson1 model extracts embedding of a fixed size (i.e., 1,024). Recordings need to be sampled at 16 kHz. Because feature extraction from long recordings required relatively complex computation, recordings were separated into 1 s segments. If a speech segment was less than 10 s, it was zero-padded to be so. For each recording, the final embedding was set to be the average of the embeddings extracted from its 10 s segments. As a pre-trained model, TRILLsson has the same advantage as x-vectors: it can be directly applied without further training. Thus, embeddings can be extracted without particular concerns about the amount of data available. To our knowledge, no previous studies have employed TRILLsson representations for the automatic detection of PD from speech.
- *Wav2Vec 2.0.* This is a self-supervised architecture that learns speech representations by masking latent representations of the raw waveform and solves a contrastive task over quantized speech representations [83]. Wav2Vec 2.0. models are widely used to extract features that can be used as input to downstream speech-related tasks, such speaker identification [84, 85], speaker verification [86], dementia, dysfluency, vocal fatigue detection, and Parkinson’s detection [87, 88, 89]. In this study, we used a ported version of S3PRL’s Wav2Vec 2.0 for the SUPERB Speaker Identification task [90]. In [90], a Wav2Vec 2.0. base-model (95*M* parameters) [83] pre-trained on Librispeech (LS-960) was used as the upstream model and fine-tuned on a Speaker Identification (SID) task^14^ using the VoxCeleb1 data set. The downstream model is constituted by mean-pooling followed by a linear transformation, and it is optimized using cross-entropy loss. Differently from the experiments in the SUPERB benchmark, in our task, we used a single-layer representation instead of the weighted sum of outputs of all layers from the base model. We used the 768*−*dimensional intermediate representations outputted from one of the layers of the model. Representations from all layers were considered. Similar to TRILLsson, because feature extraction from long recordings required relatively complex computation, long recordings were separated into 10 s segments. If a speech segment was less than 10 s, it was zero-padded to be so. At each layer, the model extracts an embedding for every 20 ms of each 10 s audio segment. A 1*−*D feature vector of size 768 representing each audio segment was then obtained by computing the mean of the embeddings along the time axis. For each recording, the final embedding was set to be the average of the feature vectors extracted from its 10 s segments. To our knowledge, we are the first to apply this representation in multi-lingual and cross-lingual PD detection.
- *HuBERT.* Among the various speech SSL models, Hidden-unit BERT (HuBERT) is one of the most prominent models for speech recognition [48]. On the LibriSpeech [91] benchmark, fine-tuned HuBERT using connectionist temporal classification (CTC) [92] achieves state-of-the-art word error rate (WER) results. Typical approaches to improve model efficiency include knowledge distillation [93], pruning [94], and model quantization [95]. Only a few works explored the performance of HuBERT on different downstream tasks. Yang et al. [90] showed that HuBERT attained the most competitive performance on a set of different downstream tasks such as speaker diarization, automatic speaker verification, and slot filling. In this study, we used a ported version of S3PRL’s HuBERT for the SUPERB SID task[90]^15^. Like Wav2Vec 2.0., we used a single-layer representation instead of the weighted sum of all layers. Representations from all layers were considered. Long recordings were separated into 10 s segments. If a speech segment was less than 10 s, it was zero-padded to be so. At each layer, the model extracts an embedding for every 20 ms of each 10 s audio segment. An 1*−*D feature vector of size 768 representing each audio segment was then obtained by computing the mean of the embeddings along the time axis. For each recording, the final embedding was set to be the average of the feature vectors extracted from its 10 s segments. To our knowledge, no previous studies have analyzed this representation for the task of PD detection.

### 3.3. Classifiers

The classification process is depicted in Figure 1 for each cross-validation iteration. The standardization part will be further explained in Section 4. As classifiers with the interpretable features, we used: Support Vector Machine (SVM), K-Nearest Neighbors (KNN), Random Forest (RF), Extreme Gradient Boosting (XGBoost), and Bagging (BG).^16^ As classifiers with non-interpretable features, we used Probabilistic Linear Discriminant Analysis (PLDA) after performing Principal Components Analysis (PCA) dimensionality reduction, similarly to Moro-Velazquez et al. [31]. The PCA transformation matrix was trained with the training embeddings and was subsequently applied to transform training, validation, and testing embeddings for each iteration. After feature dimensionality reduction to a dimensionality range between 5 to 55 with a step size of 5 by PCA (hyperparameter to be tuned), the PLDA model^17^ was trained from the PCA-reduced training subset. Both the mean of PCA-reduced PD training embeddings and the mean of PCA-reduced HC training embeddings were used as enrollment embeddings in PLDA for scoring. Given a PCA-reduced testing or validation embedding, a log-likelihood ratio with respect to the two enrollment embeddings was obtained. This ratio was compared with the equal error rate (EER) threshold from all of the ratios from the training subset. If the ratio was greater than the EER threshold, the given testing or validation sample was classified as PD. Otherwise, it was classified as HC.

**Figure 1:**
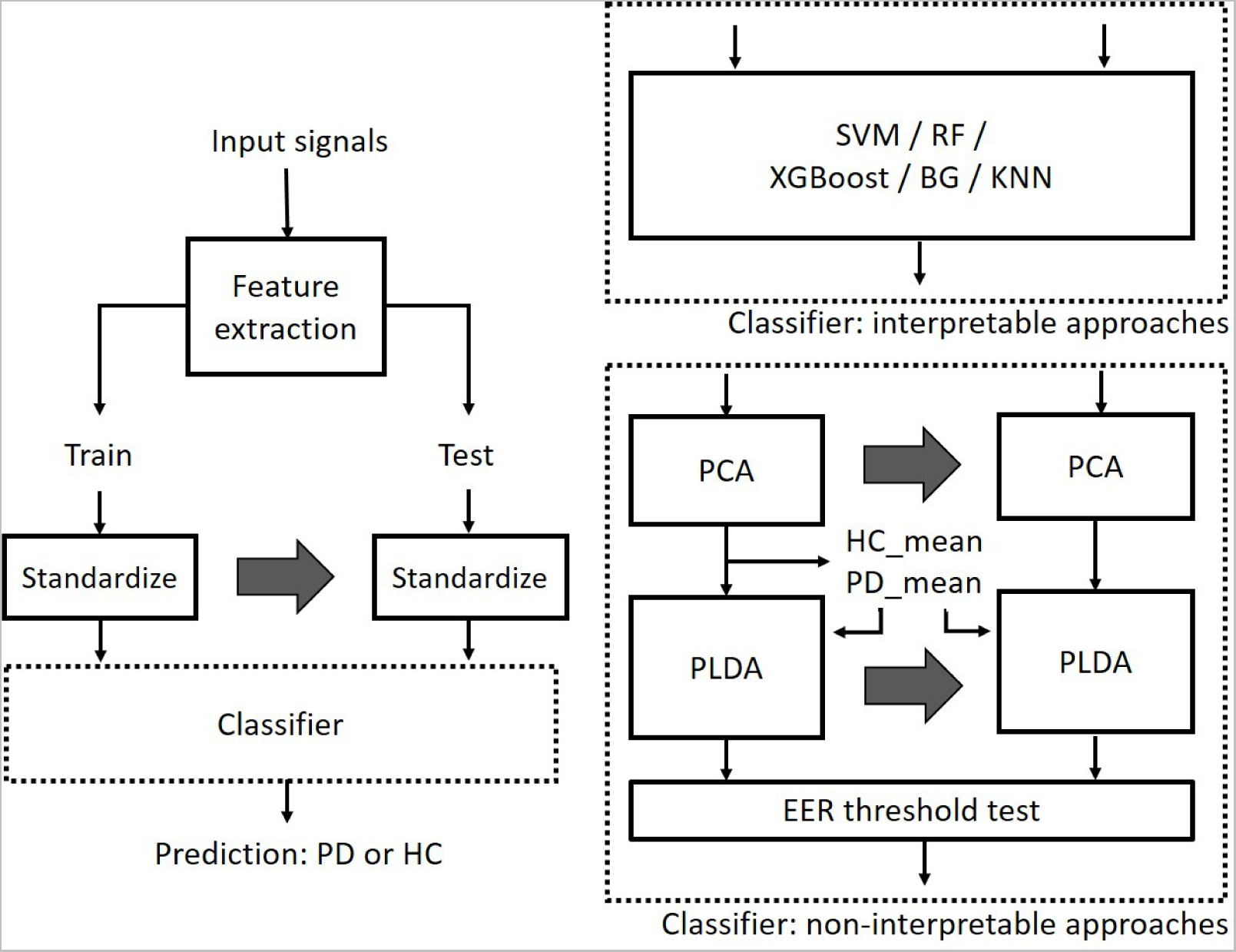
Machine learning pipeline.

### 3.4. Nested Cross-Validation (NCV)

A Nested Cross-Validation (NCV) scheme was applied in our experiments. Feature selection, hyperparameter grid search, and a train-validation-test split can improve the accuracy of ML algorithms. However, it can contribute to overfitting in most cases, especially with small data sets. NCV is a common approach that can be adopted for feature selection and parameter tuning to obtain reliable classification accuracy and prevent overfitting [96, 97]. We split the data set into ten outer folds, and each fold was held out for testing while the remaining *k −* 1 folds (9 folds) were merged to form the outer training set. Each outer training set was further split into stratified inner folds for inner training and validation. In the inner folds, we identified the 30 best features (IFMs only) and hyperparameters via grid search. An Extremely

Randomized Trees Classifier was used to perform feature selection.^18^ It is a type of ensemble learning technique which aggregates the results of multiple de-correlated decision trees to output its classification result. After getting the best hyperparameter configuration for each outer iteration using their corresponding inner folds, ten sets of unique best configurations were obtained. Because we used small data sets, the best features, and hyperparameters selected across different outer iterations could have been unstable. To evaluate more stable architectures, the standard NCV process was modified. We used the average (or mode if not applicable) configuration across the ten sets of unique best configurations as the final configuration across all outer iterations. After obtaining the final best configuration, we trained and tested the model from scratch with the final best set of features and hyperparameters on the corresponding outer training/test fold and stored the results. The final results for each model corresponded to the average model performance on the test sets of the outer folds.

We adopted the same NCV process described above in the mono-lingual and multi-lingual experiments. In doing so, we ensured that training and testing subsets never contained the same speakers. This restriction is of crucial importance since mixing the same speakers across training, and testing sets enables models to learn characteristics specific to these speakers, making final prediction results biased and over-optimistic. For the cross-lingual experiment, a direct train-test split was applied instead since testing data and training data must not come from the same language or data set. 10-fold cross-validation was then performed using all training data for hyperparameter search. Data partition details are explained in the following section.

## 4. Experiments

In this study, we employed and compared the predictive capabilities of model based on interpretable and non-interpretable representations to detect the presence of PD. All experiments were conducted task-wise, using recordings from a unique language task at a time (i.e., SS, TDU, RP). Three types of experiments were performed:

- Mono-lingual: models were trained and tested on six different data sets separately. The aim of this experiment was to compare the performance of IFMs and NIFMs.
- Multi-lingual: models were trained using training data from all the languages and tested on each of them separately. The aim of this experiment was threefold. The first was exploring the language robustness of the features used in the detection task. The second was investigating whether using more data from different languages could benefit the classification results. The third, as reported for mono-lingual experiments, was comparing the performance of IFMs and NIFMs.
- Cross-lingual: models were trained with data from all but one language used for testing. The aim of this experiment was twofold. The first was exploring the language robustness of the features used in the detection task. The second, as reported for mono-lingual and multi-lingual experiments, was comparing the performance of IFMs and NIFMs.

### 4.1. Mono-lingual

In mono-lingual experiments, NCV was applied to each data set individually, as depicted in Figure 2. Inner folds were split from the training subset assigned in outer folds. Data standardization was applied language-wise. During each outer iteration, every data set was standardized using the following transformation adapted from Kovac et al. [42]:

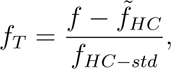

**Figure 2:**
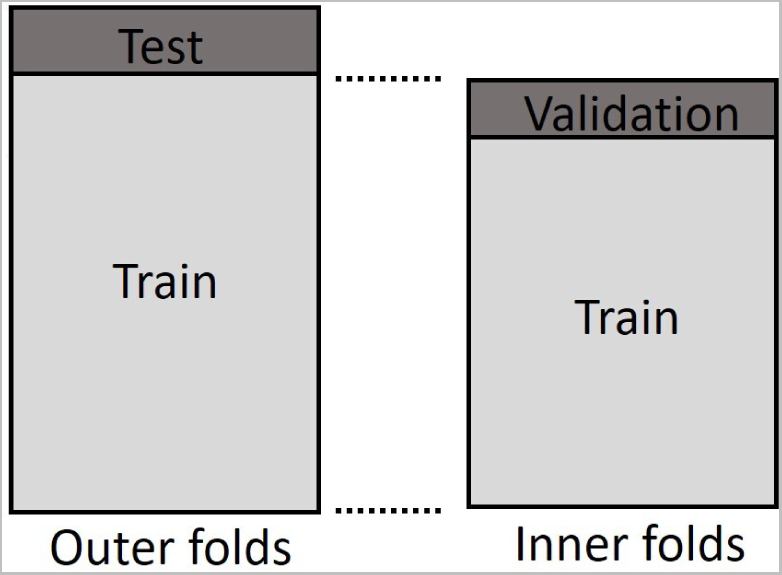
Mono-lingual experiment - data partition on a single data set.

where, for a certain feature *f*, *f_T_*is the transformed feature, 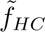 and *f_HC−std_* are the median and the standard deviation, respectively, of the HC observations for that feature in the training data.

### 4.2. Multi-lingual

In multi-lingual experiments, NCV was applied on all data sets together, as depicted in Figure 3. Each data set was split into outer folds separately. Then in each outer iteration, the training subsets of all data sets were merged into a common training set (e.g., Train 1 + Train 2 + Train 3, as shown in the figure) and split into inner folds for a grid search. After getting the optimal hyperparameters, in each outer iteration, training was conducted from scratch on the common training set, and testing was conducted on the target language test fold (e.g., Test 1, if the target data set is *data set* 1, as shown in the figure). Data standardization was performed in the same way as in mono-lingual experiments. In each outer iteration, language-wise data transformation was performed. That is, data in Train 1 and Test 1 will be standardized by the 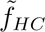 and *f_HC−std_* computed on Train 1 (see Figure 3). This type of standardization was introduced in order to avoid differences between the features’ values of the language data sets. These differences can be caused by the acoustic and linguistic peculiarities of the languages, and by the different recording conditions adopted when collecting the data.

**Figure 3:**
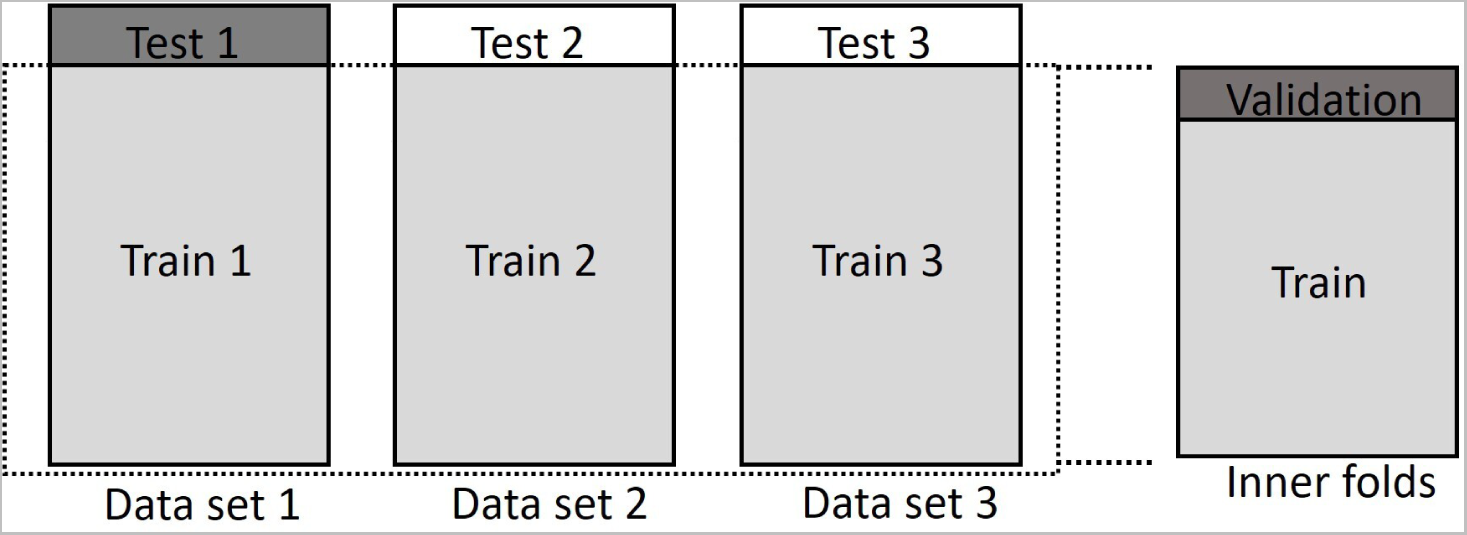
Multi-lingual experiment example - data partition on multiple data sets, with Data set 1 as the target data set to be evaluated. This illustration exemplifies our experimental approach with only three data sets but six data sets were used in our experiments.

### 4.3. Cross-lingual

The training and testing set were separated in cross-lingual experiments (see Figure 4). All data sets but the target test data set were merged into a common training set (e.g., Train 1 + Train 2) and split into cross-validation folds for a grid search. After identifying the optimized hyperparameters on the training split, training was conducted from scratch on the common training set. The testing was conducted on the target language whole data set. For training data, data standardization was applied in the same way as in mono-lingual experiments. Data transformation was performed language-wise. For testing data, since the feature distribution was unknown, features were standardized by the average 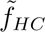 and *f_HC−std_* of all the data sets used in training. That is, data in Train 1 was standardized by 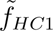 and *f_HC−std_*_1_, computed on Train 1. Data in Train 2 was standardized by 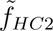 and *f_HC−std_*_2_, computed on Train 2 (see Figure 4). Lastly, data in Test was standardized by the mean of 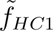 and 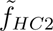 and the mean of *f_HC−std_*_1_ and *f_HC−std_*_2_.

**Figure 4:**
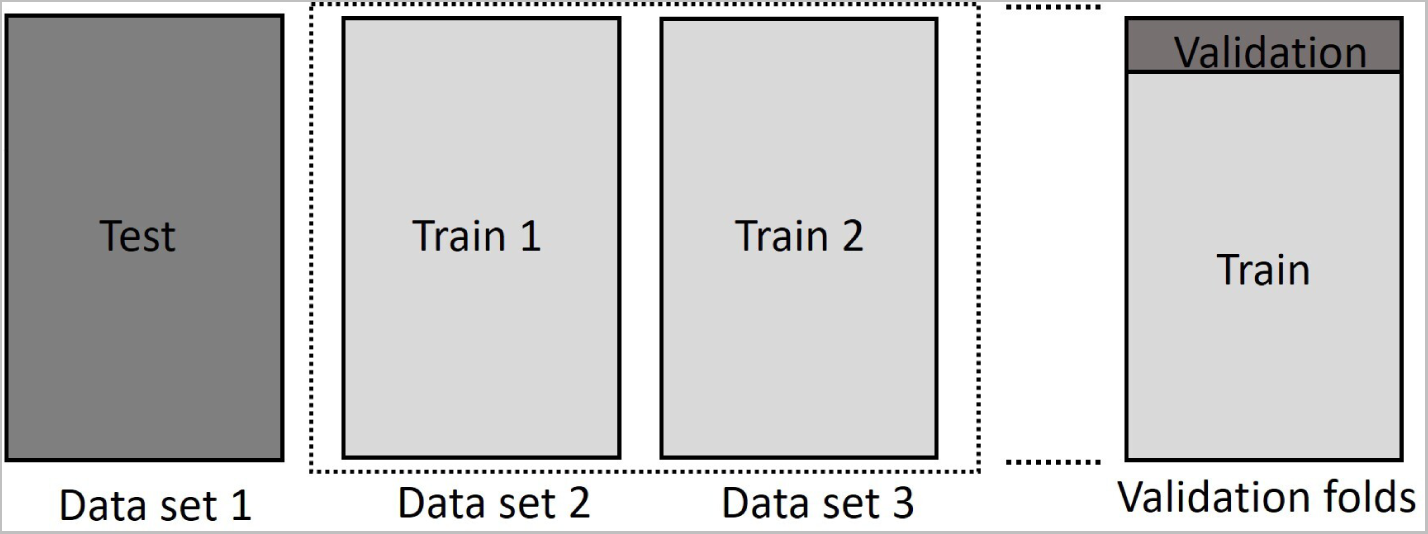
Cross-lingual experiment example - data partition on multiple data sets, with Data set 1 as the target data set to be evaluated. This illustration exemplifies our experimental approach with only three data sets but six data sets were used in our experiments.

**Figure 5:**
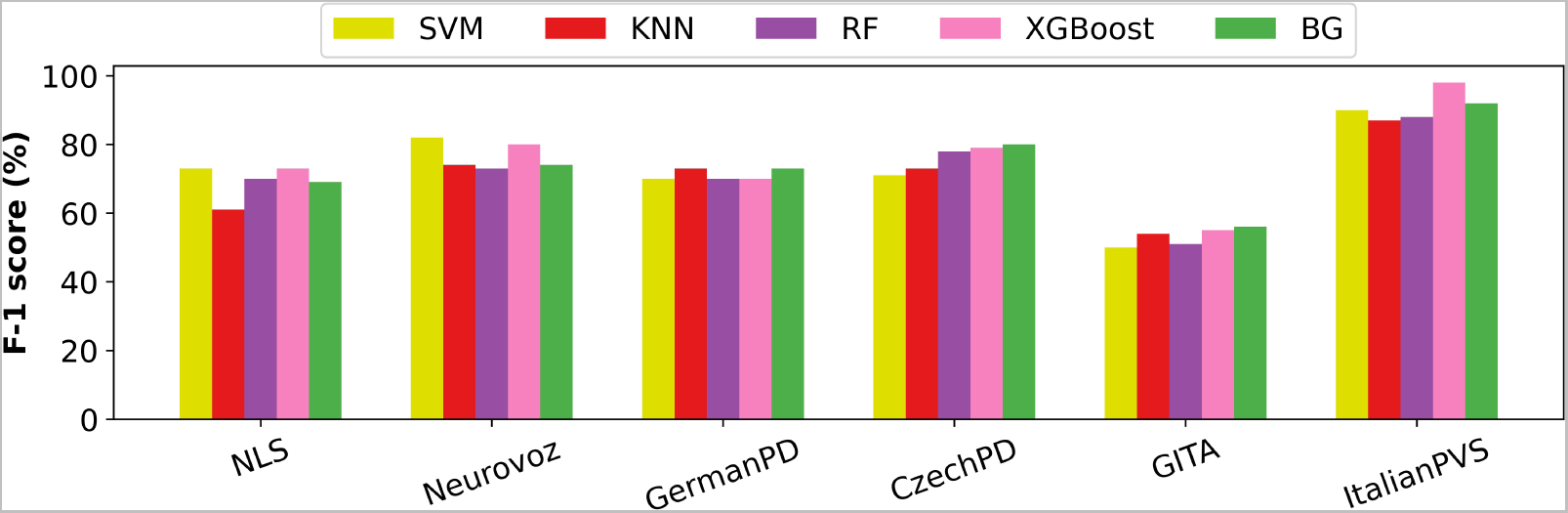
Results of mono-lingual experiments using interpretable features with 5 different classifiers: SVM (in yellow), KNN (in red), RF (in violet), XGBoost (in pink), and BG (in green). Results are reported in terms of F-1 score (%) on all the six different data sets considered. These results were obtained in the SS task, except for ItalianPVS, for which they were obtained in the TDU task.

## 5. Results and Discussion

Model performance was primarily evaluated in terms of F1-score (F1), and area under the ROC curve (AUC). Complete experimental results reporting accuracy (ACC), F1-scores, specificity (SPE), sensitivity (SEN), and AUC are reported in the Supplementary Material. For Wav2Vec 2.0. and HuBERT, all layers were experimented on. However, for Wav2Vec 2.0., embeddings extracted from the 4*^th^* layer generally performed the best. For Hubert, embeddings extracted from the 7*^th^* layer generally performed the best. Thus, only results obtained using embeddings from layer 4 (for Wav2Vec 2.0.) and layer 7 (for HuBERT) are considered. In the following subsections, we first discuss the results for IFMs and NIFMs separately. We then compare their performances in mono-lingual, multi-lingual, and cross-lingual experiments, respectively. The differences in model performances are reported in absolute terms.

### 5.1. Interpretable features

In mono-lingual experiments, F1-scores ranged from 50% to 98% and AUC from 0.50 to 1.00. In multi-lingual experiments, F1-scores ranged from 50% to 87%, and AUC from 0.56 to 0.93. In cross-lingual experiments instead, performance significantly varied depending on the target language used in testing. In some experiments, higher performance was achieved using SS samples, while in others read speech samples. Figure 6 shows the results reported by the different classifiers employed during the mono-lingual experiments. Table 5 instead reports results obtained in mono-lingual, multilingual, and cross-lingual experiments when employing XGBoost as classifier. In this regard, XGBoost performed better on average than the other types of classifiers across data sets and tasks. ItalianPVS and Neurovoz were the two data sets on which IFMs achieved the best results in terms of F1 and AUC.

**Figure 6:**
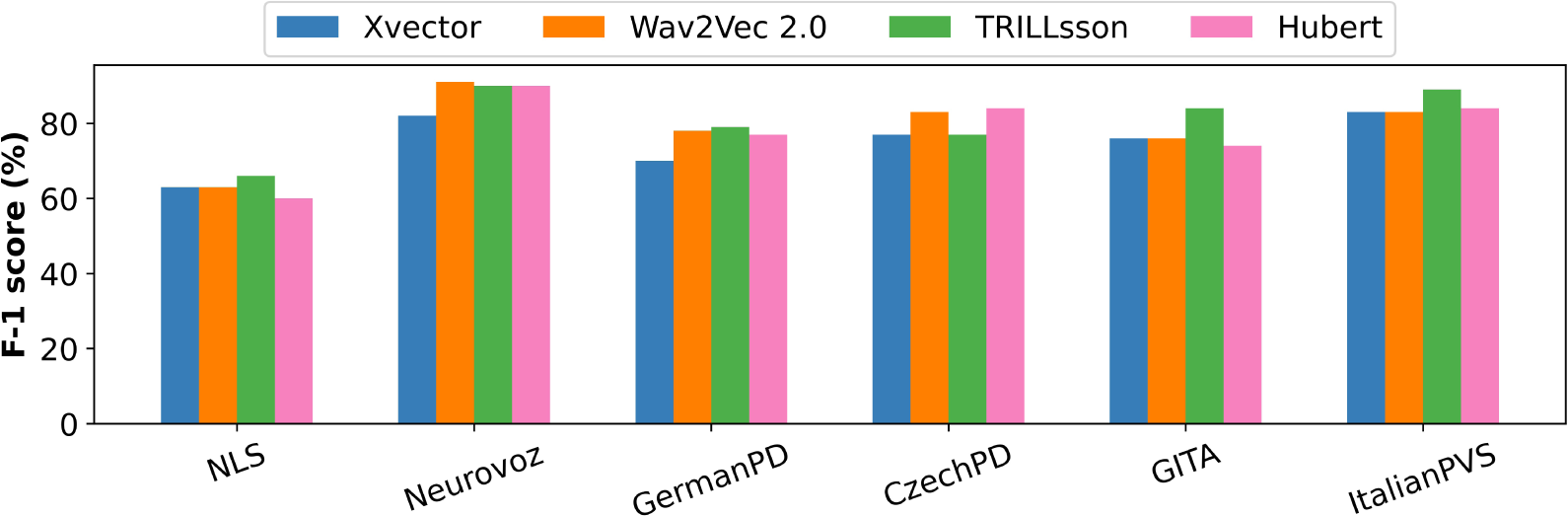
Results of mono-lingual experiments using non-interpretable features: x-vectors (in blue), Wav2Vec 2.0 (in orange), TRILLsson (in green) and Hubert (in pink). Results are reported in terms of F-1 score (%) on all the six different data sets considered. These results were obtained in the RP task, except for Neurovoz, for which they were obtained in the TDU task.

**Table 5:** Results of mono-lingual, multi-lingual, and cross-lingual experiments using interpretable features and XGBoost as classifier. Results are reported for SS, RP, and TDU tasks respectively. F-1 score and area under the ROC (AUC) are used as evaluation metrics. Abbreviations: mono, mono-lingual; multi, multi-lingual; cross, cross-lingual.

|  |  | Mono |  | Multi |  | Cross |  |
| --- | --- | --- | --- | --- | --- | --- | --- |
| Test | Task | F1 | AUC | F1 | AUC | F1 | AUC |
| NLS<br>(American English) | SS | 0.73 | 0.80 | 0.70 | 0.77 | 0.72 | 0.75 |
|  | RP | 0.60 | 0.63 | 0.72 | 0.77 | 0.72 | 0.79 |
| Neurovoz<br>(Castilian Spanish) | SS | 0.80 | 0.93 | 0.80 | 0.80 | 0.69 | 0.71 |
|  | TDU | 0.71 | 0.88 | 0.74 | 0.86 | 0.68 | 0.79 |
| GermanPD<br>(German) | SS | 0.70 | 0.71 | 0.71 | 0.75 | 0.52 | 0.60 |
|  | RP | 0.71 | 0.77 | 0.76 | 0.83 | 0.53 | 0.60 |
| CzechPD<br>(Czech) | SS | 0.79 | 0.70 | 0.82 | 0.65 | 0.49 | 0.69 |
|  | RP | 0.71 | 0.85 | 0.72 | 0.80 | 0.70 | 0.79 |
| GITA<br>(Colombian Spanish) | SS | 0.55 | 0.57 | 0.60 | 0.62 | 0.51 | 0.55 |
|  | RP | 0.70 | 0.71 | 0.66 | 0.74 | 0.51 | 0.50 |
| ItalianPVS<br>(Italian) | RP | 0.92 | 0.98 | 0.85 | 0.88 | 0.77 | 0.76 |
|  | TDU | 0.98 | 1.00 | 0.86 | 0.87 | 0.64 | 0.83 |

### 5.2. Non Interpretable features

In mono-lingual experiments, F1-scores ranged from 63% to 91% and AUC from 0.64 to 1.00. In multi-lingual experiments, F1-scores ranged from 62% to 95% and AUC from 0.71 to 0.99. Similarly to what was observed for IFMs, in cross-lingual experiments, the performance of NIFMs did significantly vary depending on the target language considered in the experiments. Overall, TRILLsson, Wav2Vec 2.0., and HuBERT reported better classification results than x-vectors in mono-lingual and multi-lingual experiments. Experimental results obtained with non-interpretable features in the RP task and the TDU task (for Neurovoz only), respectively, are displayed in Fig. 6 (mono-lingual), 7 (multi-lingual), and 8 (cross-lingual).

**Figure 7:**
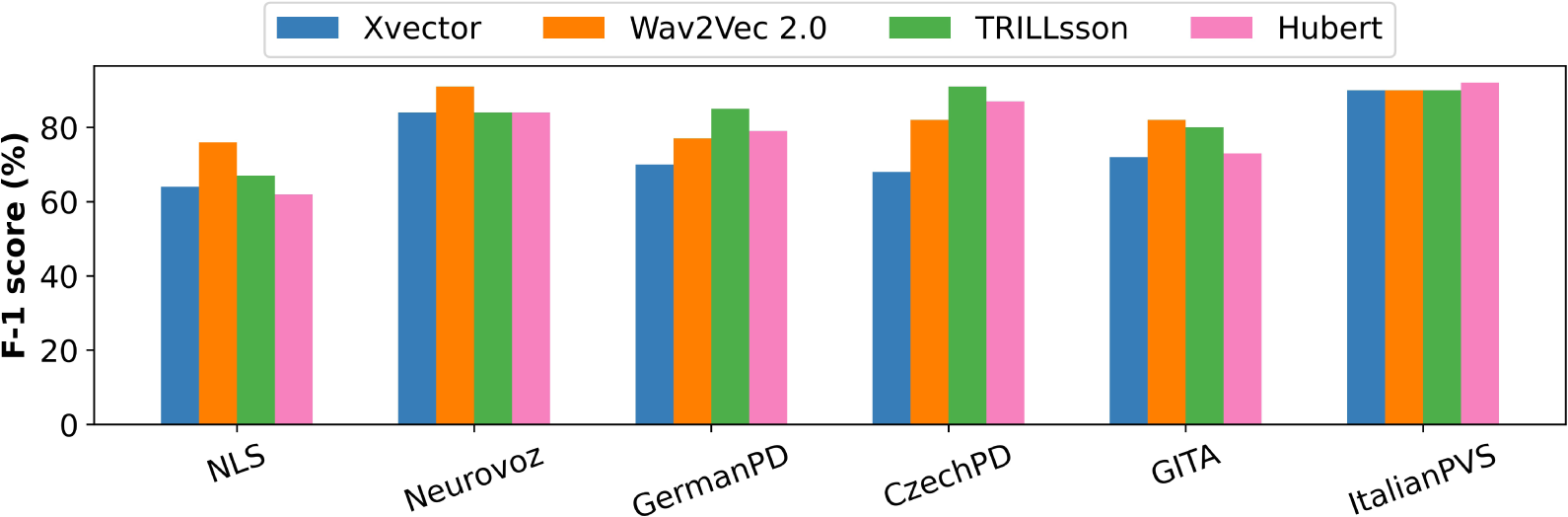
Results of multi-lingual experiments using non-interpretable features: x-vectors (in blue), Wav2Vec 2.0 (in orange), TRILLsson (in green) and Hubert (in pink). Results are reported in terms of F-1 score (%) on all the six different data sets considered. These results were obtained in the RP task, except for Neurovoz, for which they were obtained in the TDU task.

**Figure 8:**
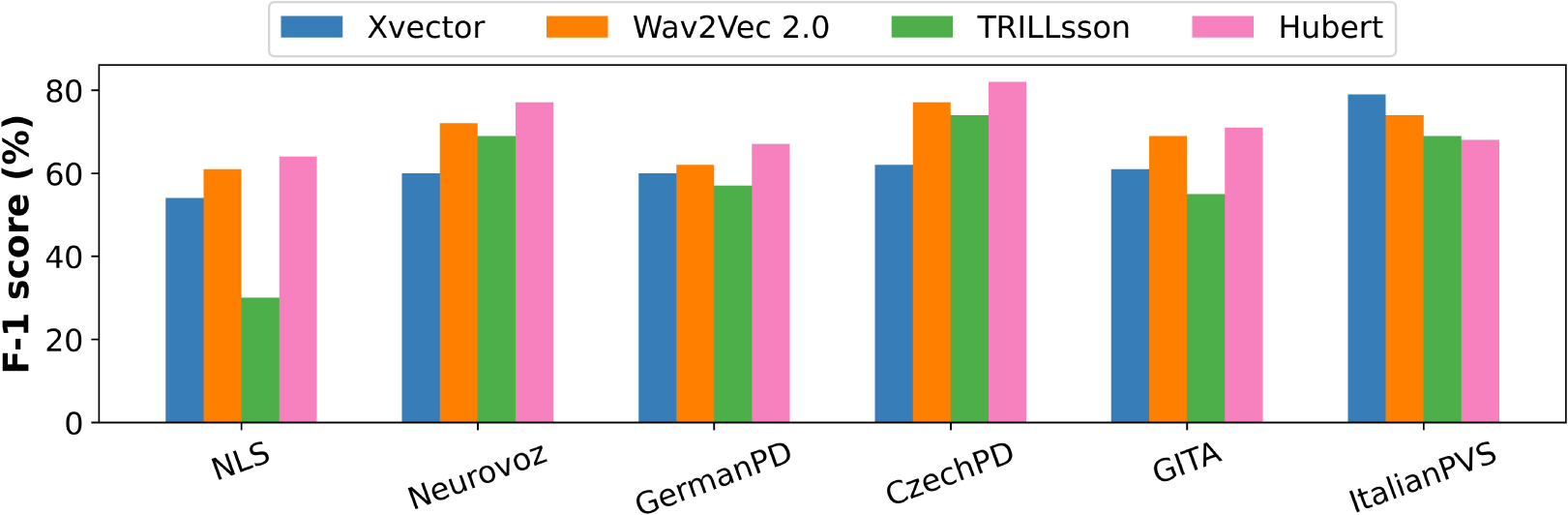
Results of cross-lingual experiments using non-interpretable features: x-vectors (in blue), Wav2Vec 2.0 (in orange), TRILLsson (in green) and Hubert (in pink). Results are reported in terms of F-1 score (%) on all the six different data sets considered. These results were obtained in the RP task, except for Neurovoz, for which they were obtained in the TDU task.

### 5.3. Interpretable vs. Non-Interpretable Features

Figures 9 and 10 report the best experimental results in terms of F1-score and AUC obtained in mono-lingual, multi-lingual, and cross-lingual experiments using interpretable and non-interpretable features, respectively. Results are reported language-wise for each task analyzed.

**Figure 9:**
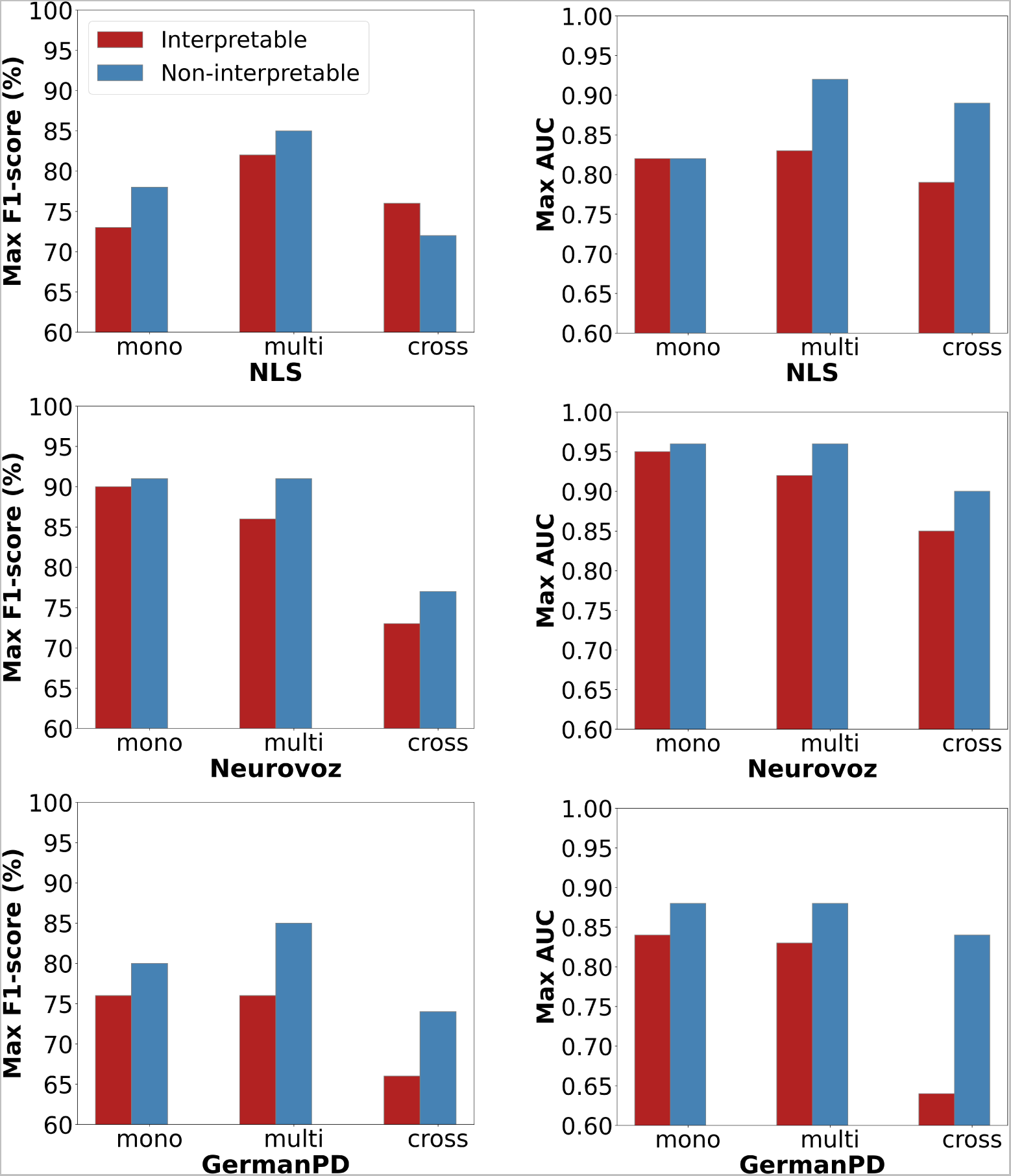
Bar plots describing the best F1-scores (left panel) and AUC (right panel) across tasks using IFMs (in red) and NIFMs (in blue), respectively. For each evaluation metric, mono-lingual, multi-lingual, and cross-lingual experimental results on NLS, Neurovoz, and GermanPD are reported. Abbreviations: Mono, Mono-lingual; Multi, Multi-lingual; Cross, Cross-lingual.

**Figure 10:**
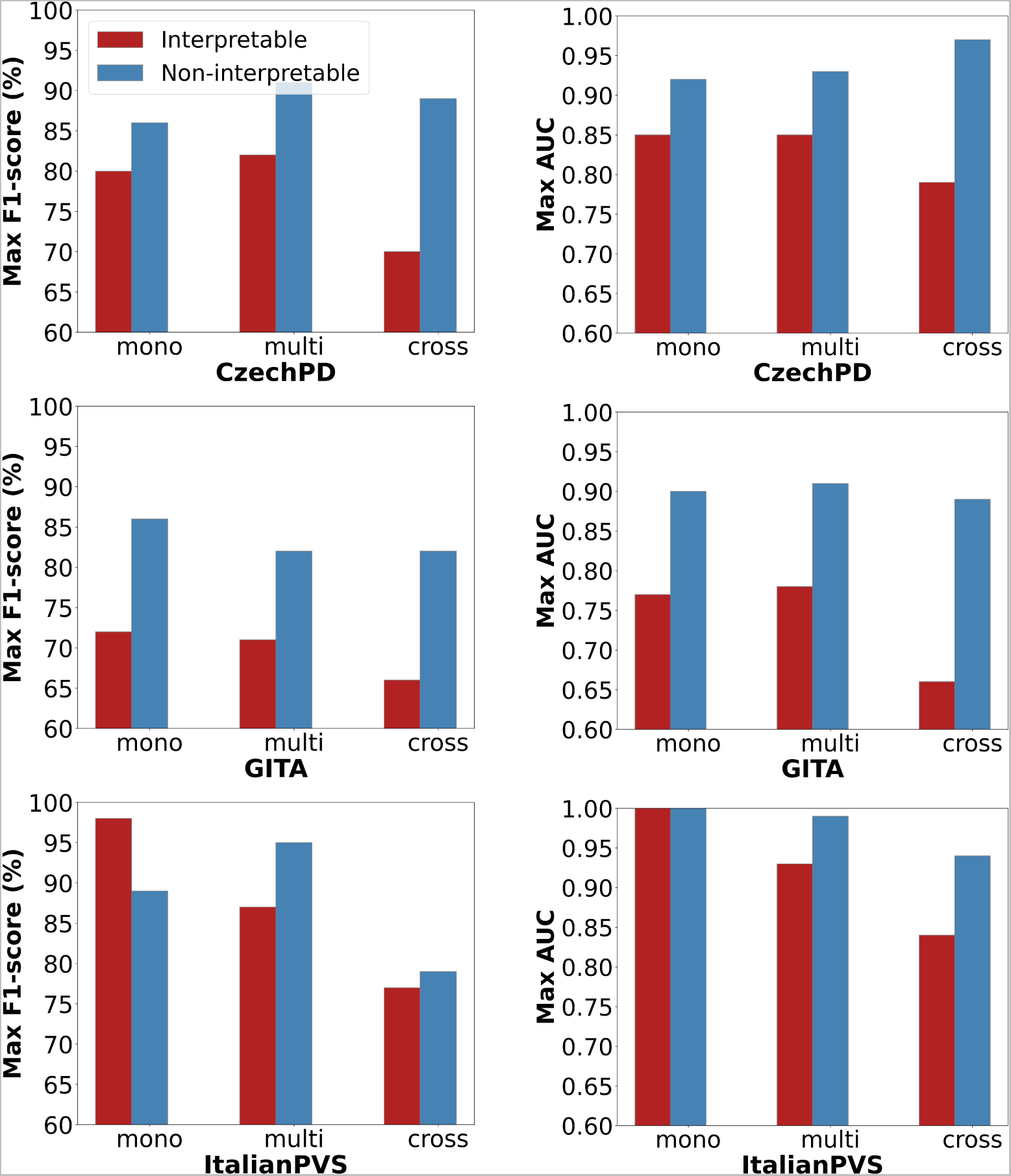
Bar plots describing the best F1-scores (left panel) and AUC (right panel) across tasks using IFMs (in red) and NIFMs (in blue), respectively. For each evaluation metric, mono-lingual, multi-lingual, and cross-lingual experimental results on CzechPD, GITA, and ItalianPVS are reported. Abbreviations: Mono, Mono-lingual; Multi, Multi-lingual; Cross, Cross-lingual.

#### Mono-lingual

For IFMs, the mean of the best F1-scores across languages was 81%, and that of AUC was 0.87. For NIFMs, the mean of best F1-scores across languages was 85%, and that of AUC was 0.91. In particular, NIFMs worked better than IFMs in Colombian Spanish, Czech, and German. On Italian, Castilian, Spanish, and American English, IFMs achieved similar results to those reported by NIFMs and, in some cases, slightly better, as observed on ItalianPVS. On average NIFMs performed better than IFMs by *∼*4% and 0.04 considering F1-score and AUC, respectively.

In mono-lingual experiments, NIFMs had more inconsistent distributions in specific languages, resulting in varying optimal decision thresholds. This fact contributed to a discrepancy between F1 and AUC results. The classification results achieved using interpretable features were lower than those achieved using non-interpretable features. However, our IFMs reported an improvement over previous models. Namely, the performances of our best IFMs on CzechPD and ItalianPVS outperformed those reported by Kovac et al. [41, 42] based on acoustic descriptors by more than 10 percentual absolute points in ACC. Similarly, TRILLsson reported best AUCs of 0.95, 0.87, 0.84 on Neurovoz, GITA, and GermanPD, which were 0.01, 0.03, and 0.16 higher than the x-vector approach of [31], the CNN approach of [28], and the mono-lingual CNN approach of [27], respectively. In this regard, HuBERT/Wav2Vec 2.0. reported best AUCs of 0.96, 0.90, and 0.88 on Neurovoz, GITA, and GermanPD, respectively, which were even higher than those achieved with TRILLsson.

#### Multi-lingual

For IFMs, the mean of the best F1-scores across each language was 81%, and that of AUC was 0.86. For NIFMs, the mean of the best F1-scores across each language was 88%, and that of AUC was 0.93. On average NIFMs performed better than IFMs by 7% and 0.07 considering F1-score and AUC, respectively.

For both IFMs and NIFMs, using a multi-lingual training data set led to an improvement *≥*0.05 for both F1 and AUC in some tasks in almost all languages, with respect to the mono-lingual experiments. In addition, universal paralinguistic representations provided by TRILLsson allowed reaching better performances compared to x-vectors and other non-interpretable representations. In a previous study, Vásquez-Correa et al. [27] increased the AUC on the test set of GermanPD from 0.68 to 0.82 by adding GITA in the training set, and the test AUC on GITA from 0.78 to 0.82 by adding Czech data in the training set. Similarly, our TRILLsson-based models increased the AUC on the test set of GermanPD (RP task) from 0.83 to 0.88 and that on GITA (SS task) from 0.84 to 0.91 in the multi-lingual scenario. Altogether, the results of multi-lingual experiments confirmed some level of language robustness of all the representations adopted.

#### Cross-lingual

In cross-lingual experiments, performance significantly varied depending on the target language considered in the experiments. For IFMs, the mean of best F1-scores across each language was 71%, and that for AUC was 0.76. For NIFMs, the mean of best F1-scores across each language was 79%, and that for AUC was 0.91. Even though results achieved using interpretable features were lower than those reported using non-interpretable ones, our IFMs reported an improvement over previous IFMs reported in the literature that adopted a similar training scheme (i.e., leave-one-language-out). Namely, our best IFMs on CzechPD and GITA outperformed those of Kovac et al. [42] by 12% in terms of ACC. Our models reported higher SEN and SPE by approximately 10% for American English and 5% higher for ItalianPVS than than those reported by Kovac et al. [41].

For both IFMs and NIFMs, we observed a performance improvement *≥*0.05 in F1-score and AUC with respect to mono-lingual experiments in only a few data sets (i.e., NLS for IFMs and HuBERT-based models, and CzechPD for TRILLsson based models). In most cases, classifying subjects from a language group not included in the training set led to a significant decrease in the classification results, as shown in previous works [41, 42, 40]. In addition, not seeing the test language in the training set might have contributed to non-optimal data standardization and shifted distributions of classifier decision scores, making F1-scores unstable, especially for NIFMs.

## 6. Additional Remarks

In this section, we analyze the limitations observed in the present work. The data sets adopted differ in size and case severity. Thus, in multi-lingual and cross-lingual experiments, the training and testing sets might not be severity-matched. The time since the last medication, as well as the peak-dose duration, may impact performance. In some studies, participants with PD received medication before starting the recording session, which can result in better speech and language performances. In contrast, they did not receive any medication in other data sets (e.g., CzechPD). Furthermore, even though we analyzed three different types of tasks (i.e., TDU, RP, SS), only some data sets contain all of them. This fact limits the generalizability of our results and represents a limitation in multi-lingual and cross-lingual experiments. In addition, even though we performed the experiments task-wise, comparing the predictive capabilities of models based on features extracted from the same type of tasks, there could be differences occurring between variants of the same task. These differences can influence the feature values and make the comparison between languages biased. More generally, if a data set has relatively more complex tasks than others in terms of cognitive load or tasks’ length, this fact might affect the features extracted and, in turn, the classification results. Finally, even though the same machine learning pipeline has been followed to perform the experiments using interpretable and non-interpretable features, we are not using the same classifiers for the two types of representations. This fact could have affected the models’ performances as well. On one hand, classifiers like SVM, random forest, and bagging are used with low-dimensional data. On the other, PCA and PLDA techniques are typically employed with speech embeddings because they are better suited for high-dimensional data and can provide better generalization performance [72].

## 7. Conclusions and Future Work

The proposed study presents a systematic comparison between interpretable and non-interpretable speech-based representations for the automatic detection of PD. Six data sets containing speech recordings from different languages were used, which allows comparison in mono-lingual, multi-lingual, and cross-lingual experiments. The set of interpretable features encompassed prosodic descriptors (e.g., pitch, loudness variation, and speech pauses), linguistic descriptors (e.g., frequency of occurrence of different POS and syntactic complexity), and cognitive descriptors (e.g., regularity of speech rhythm and the number of correct IU). The set of non-interpretable features included x-vectors, TRILLsson, Wav2Vec 2.0., and HuBERT speech representations, with TRILLsson and HuBERT being applied to PD detection for the first time. Based on the experimental results, we observed that:

1. In mono-lingual experiments, where classifiers were trained and tested on each data set separately, non-interpretable representations outperformed interpretable ones on average by *∼*4% and 0.04 considering F1-score and AUC, respectively. In this regard, HuBERT, Wav2Vec 2.0., and TRILLsson provided better and much more stable results compared to x-vectors.
2. In multi-lingual experiments, where classifiers were trained on all languages and tested on each language individually, NIFMs performed better than IFMs by *∼*7% and 0.07 considering F1-score and AUC, respectively, with Wav2Vec 2.0. and TRILLsson reporting the best performances. Both approaches reported a performance improvement *≥*0.05 with respect to mono-lingual results in some tasks across almost all languages. This suggests that using more data, even when it comes from other languages, helps improve classification results. This phenomenon can be motivated by the fact that there are common patterns in PD-related dysarthria across languages that can be leveraged to train more robust models.
3. In cross-lingual experiments, where classifiers were trained on all languages but the target test language, NIFMs outperformed IFMs by *∼*5.8% and 0.14 in terms of F1-score and AUC, respectively, with TRILLsson reporting the best performance. A performance improvement *≥*0.05 in AUC with respect to mono-lingual results was observed in only a few data sets (i.e., NLS for IFMs and HuBERT-based models, and CzechPD for TRILLsson-based models).

Some more general considerations are worth mentioning. First, using TRILLsson, Wav2Vec 2.0., and HuBERT self-supervised representations reported better classification results than using x-vectors in mono-lingual and multi-lingual experiments. Using heavier pre-trained models that are more complex in architecture, such as TRILLsson 2*∼*5 or larger Wave2Vec 2.0. and HuBERT models, might further increase the performance too. Second, in cross-lingual experiments, TRILLsson significantly outperformed x-vectors and reported a better performance compared to Wav2Vec 2.0. and HuBERT in terms of AUC but, in some cases, performed very poorly in terms of F1-score.

Overall, the performance of NIFMs was significantly better than that of IFMs, especially in cross-lingual scenarios, even though the difference in the performance between IFMs and NIFMs was not always remarkable, as shown in some mono-lingual and multi-lingual experiments. Altogether, the results of these experiments did not confirm the findings reported in previous studies that suggested that providing a structured data set with a comprehensive representation of meaningful features can considerably enhance classification results and narrow the performance gap between IFMs and NIFMs [25, 45, 44, 98, 99, 100]. In the current work, paralinguistic representations from DNNs seem to deliver a more exhaustive characterization of the pathological speech of PD individuals. However, the performance of models based on these complex features can still be complemented with hand-crafted features. Hence, clinicians can leverage IFMs to have clear insights into the evolution and the possible deterioration of the spoken production of patients with PD, while NIFMs can be employed to achieve higher detection accuracy.

In future work, we intend to validate the experimental results obtained in this work, balancing the classes in terms of the number of subjects, age, gender, time of medication, PD disease severity, and specific phenotype. We also plan to explore the language robustness of a more comprehensive set of interpretable and non-interpretable speech-based representations to provide a more exhaustive characterization of PD and to experiment with new classification techniques. Finally, as individuals with Parkinson’s disease mimics conditions might be often misdiagnosed with PD, it is important to create a separate group when performing differential diagnosis. Altogether, both IFMs and NIFMs showed a satisfying generalization capability in multi-lingual and cross-lingual experiments. As such, after further validation in clinical trials, they might be used to deploy screening tools for the automatic assessment of speech of people with PD in different languages.

## Data Availability

Neurological Signals was collected by the authors of this study. Data are protected by privacy and security law. The authors may share this data set with the public once the data collection is completed. This is allowed by the license and the agreement signed by participants. Neurovoz, GITA,  GermanPD, and CzechPD are not publicly accessible. The authors of these data sets may or may not want to share their data with the public. ItalianPVS can be found at the following link reported below. 

https://ieee-dataport.org/open-access/italian-parkinsons-voice-and-speech

## 8. Abbreviations

PD: Parkinson’s Disease
HC: Healthy Control
DL: Deep Learning
ML: Machine Learning
CNN: Convolutional Neural Network
MFCC: Mel-Frequency Cepstral Coefficient
ASR: Automatic Speech Recognition
POS: Part of Speech
IU: Informational Unit
SVM: Support Vector Machines
KNN: K-Nearest Neighbors
RF: Random Forest
XGBoost: Extreme Gradient Boosting
BG: Bagging
PCA: Principal Components Analysis
PLDA: Probabilistic Linear Discriminant Analysis
EER: Equal Error Rate
NCV: Nested Cross-Validation
ROC: Receiver Operating Characteristic
AUC: Area under the Receiver Operating Characteristic curve
SS: Spontaneous Speech
RP: Reading Passage
TDU: Text-Dependent Utterances
ACC: Accuracy
SEN: Sensitivity
SPE: Specificity
IM: Interpretable Feature-based Model
NIFM: Non-Interpretable Feature-based Model
Mono: Mono-lingual
Multi: Multi-lingual
Cross: Cross-lingual.

## 9. Conflict of Interest Statement

We have no conflicts of interest to disclose.

## 10. Author Contributions

AF contributed to conceptualization, data pre-processing, features extraction, machine learning, GitHub repository management, and article writing. YT contributed to conceptualization, feature extraction, machine learning, GitHub repository management, and article writing. LMV contributed to conceptualization, data collection, project supervision, and article writing. AB and ND contributed to data collection, conceptualization, project supervision, and manuscript revision. TT and JV contributed to conceptualization, project supervision, and manuscript revision. All authors contributed to the manuscript revision, read, and approved the submitted version.

## 11. Funding

This work was funded in part by the Richman Family Precision Medicine Center of Excellence – Venture Discovery Fund and Consolidated Anti-Aging Foundation.

## 12. Data Availability Statement

NLS was collected by the authors of this study. Data are protected by privacy and security law. The authors may share this data set with the public once the data collection is completed. This is allowed by the license and the agreement signed by participants. Neurovoz, GITA, GermanPD, and CzechPD are not publicly accessible. The authors of these data sets may or may not want to share their data with the public. ItalianPVS can be found at the following link: https://ieee-dataport.org/open-access/italian-p arkinsons-voice-and-speech. Information for this data set is contained within the article [101]. The code to reproduce our experiments can be found at the following link: https://github.com/Neuro-Logical/speech/tree/main/Cross_Lingual_Evaluation.

## Acknowledgements

None.

## Footnotes

1 https://ieee-dataport.org/open-access/italian-parkinsons-voice-and-speech (last accessed on 4 February 2023)

2 https://sox.sourceforge.net/, last accessed on 26 January 2023.

3 https://pypi.org/project/ffmpeg-normalize/, last accessed on 26 January 2023.

4 https://openai.com/blog/whisper/

5 https://github.com/jcvasquezc/DisVoice

6 https://parselmouth.readthedocs.io/en/stable/

7 https://www.google.com/search?q=praat+software&oq=praat+software&aqs=chrome.0.0i512l2j0i22i30l8.4071j0j15&sourceid=chrome&ie=UTF-8

8 https://github.com/NeuroLexDiagnostics/DigiPsych_Prosody

9 https://github.com/wiseman/py-webrtcvad

10 https://spacy.io/models

11 The code used to extract word token timestamp is available at https://github.com/jianfch/stable-ts.

12 https://huggingface.co/speechbrain/spkrec-xvect-voxceleb

13 https://tfhub.dev/s?q=trillsson

14 https://huggingface.co/superb/wav2vec2-base-superb-sid

15 https://huggingface.co/superb/HuBERT-base-superb-sid

16 To implement the different classifiers, we used the Scikit-learn (Sklearn) library in Python. See https://www.tutorialspoint.com/scikit_learn/scikit_learn_introduction.htm

17 The PLDA model was implemented by the SpeechBrain toolkit. See https://speechbrain.readthedocs.io/en/latest/API/speechbrain.processing.html

18 The documentation of the Python library used to implement the Extremely Randomized Trees Classifier can be found at https://scikit-learn.org/stable/modules/generated/sklearn.ensemble.ExtraTreesClassifier.html.

## Notes

### Competing Interest Statement

The authors have declared no competing interest.

### Author Declarations

The authors of this study collected a data set called NeuroLogical Signals (NLS) at Johns Hopkins Medicine (JHM). The participants were categorized as either having a neurological disorder or being healthy controls and received treatment and diagnosis from expert neurologists at JHM. All participants underwent informed consent, and the data collection was approved by the Johns Hopkins Medical Institutional Review Board.

