## Supplemental Tables 1, 2, and 3. for "Interpretable Speech Features vs. DNN Embeddings: What to Use in the Automatic Assessment of Parkinson’s Disease in Multi-lingual Scenarios"

### Supplementary Material

#### 1. Supplementary Tables

| **Interpretable Features** | | | |
|---|---|---|---|
| **Feature family** | **Feature name** | **Feature description** | **Library** |
| Prosody | Features based on F0 | 1-6 F0-contour Avg., Std., Max., Min., Skew., Kurt<br>7-12 Tilt of a linear estimation of F0 for each voiced segment Avg., Std., Max., Min., Skew., Kurt.<br>13-18 MSE of a linear estimation of F0 for each voiced segment Avg., Std., Max., Min., Skew., Kurt.<br>19-24 F0 on the first voiced segment Avg., Std., Max., Min., Skew., Kurt.<br>25-30 F0 on the last voiced segment Avg., Std., Max., Min., Skew., Kurt. | Disvoice |
| | Features based on energy | 31-34 energy-contour for voiced segments Avg., Std., Skew., Kurt.<br>35-38 Tilt of a linear estimation of energy contour for V segments Avg., Std., Skew., Kurt.<br>39-42 MSE of a linear estimation of energy contour for V segment Avg., Std., Skew., Kurt.<br>43-48 energy on the first voiced segment Avg., Std., Max., Min., Skew., Kurt.<br>49-54 energy on the last voiced segment Avg., Std., Max., Min., Skew., Kurt.<br>55-58 energy-contour for unvoiced segments Avg., Std., Skew., Kurt.<br>59-62 Tilt of a linear estimation of energy contour for U segments Avg., Std., Skew., Kurt.<br>63-66 MSE of a linear estimation of energy contour for U segments Avg., Std., Skew., Kurt.<br>67-72 energy on the first unvoiced segment Avg., Std., Max., Min., Skew., Kurt.<br>73-78 energy on the last unvoiced segment Avg., Std., Max., Min., Skew., Kurt. | |
| | Features based on duration | 79 Voiced rate Number of voiced segments per second<br>80-85 Duration of Voiced Avg., Std., Max., Min., Skew., Kurt<br>86-91 Duration of Unvoiced Avg., Std., Max., Min., Skew., Kurt.<br>92-97 Duration of Pauses Avg., Std., Max., Min., Skew., Kurt.<br>98-103 Duration ratios Pause/(Voiced+Unvoiced), Pause/Unvoiced,<br>Unvoiced/(Voiced+Unvoiced), Voiced/(Voiced+Unvoiced),<br>Voiced/Pause, Unvoiced/Pause | |
| | Features based on loudness | 104 Loudness variability | Parselmouth |
| | Features based on pauses | 105-107 Total Time<br>108-110 Total Speech Time<br>111-113 Total Pause Time<br>114-116 Percentage Pause Time<br>117-119 Speech Pause Time<br>120-122 Mean Pause Duration<br>123-125 Pause Variability | DigiPsych Prosody |
| Linguistic | Feature based on part-of-speech (POS) | 1-10 Word count, word count with no functional words, avg. word length,<br>sentence count, avg. sentence length in words, noun count, verb count,<br>numeral count, auxiliary count, adjective count, | Whisper + Spacy |
| | Features based on syntactic phrases | 11-13 Number of noun phrases, number of verb phrases,<br>number of prepositional phrases | |
| Cognitive | Features based on speech rhythm | 1-3 Feature based on speech rhythm Std., Skew., Kurt. | Whisper |
| | Features based on IUs | 4 Number of correct informational verbs | Whisper + Spacy |

Table 1: Summary of the family of interpretable features extracted. For each feature family, we report the name of the features, with the number and description of the coefficients extracted and the library employed for the extraction. Abbreviations: Avg., Average; Std., Standard Deviation; Max, Maximum; Min, Minimum; Skew, Skewness; Kurt, Kurtosis.

| **Mono-lingual** | | | | | | | | | | | | | | | | | | | | | | | | | | | |
|---|---|---|---|---|---|---|---|---|---|---|---|---|---|---|---|---|---|---|---|---|---|---|---|---|---|---|---|
| Test | Task | spk# | SVM | | | | | KNN | | | | | Random Forest | | | | | XGBoost | | | | | Bagging | | | | |
| | | | ACC | F1 | SPE | SEN | AUC | ACC | F1 | SPE | SEN | AUC | ACC | F1 | SPE | SEN | AUC | ACC | F1 | SPE | SEN | AUC | ACC | F1 | SPE | SEN | AUC |
| NLS | SS | PD-23, HC-27 | 0.76 | **0.73** | 0.69 | 0.84 | **0.82** | 0.63 | 0.61 | 0.53 | 0.73 | 0.57 | 0.70 | 0.70 | 0.67 | 0.73 | 0.80 | 0.74 | **0.73** | 0.62 | 0.73 | 0.80 | 0.68 | 0.69 | 0.65 | 0.73 | 0.78 |
| | RP | PD-23, HC-27 | 0.66 | 0.62 | 0.58 | 0.65 | 0.60 | 0.70 | **0.70** | 0.53 | 0.78 | **0.68** | 0.62 | 0.50 | 0.58 | 0.75 | 0.63 | 0.62 | 0.60 | 0.53 | 0.72 | 0.63 | 0.68 | 0.68 | 0.58 | 0.72 | 0.61 |
| Neurovoz | TDU | PD-43, HC-46 | 0.82 | 0.77 | 0.85 | 1.00 | 0.92 | 0.83 | 0.83 | 0.86 | 1.00 | 0.92 | 0.88 | **0.90** | 0.96 | 0.96 | **0.95** | 0.82 | 0.80 | 0.96 | 0.94 | 0.93 | 0.86 | 0.86 | 0.96 | 0.94 | 0.94 |
| | SS | PD-20, HC-21 | 0.85 | **0.82** | 0.75 | 0.85 | 0.82 | 0.74 | 0.74 | 0.45 | 0.85 | **0.83** | 0.76 | 0.73 | 0.70 | 0.70 | 0.81 | 0.71 | 0.71 | 0.65 | 0.80 | 0.88 | 0.74 | 0.74 | 0.70 | 0.80 | 0.88 |
| German | SS | PD-88, HC-88 | 0.57 | 0.70 | 1.00 | 0.08 | 0.57 | 0.73 | 0.73 | 0.77 | 0.63 | 0.75 | 0.69 | 0.70 | 0.73 | 0.68 | 0.72 | 0.69 | 0.70 | 0.73 | 0.66 | 0.71 | 0.73 | **0.73** | 0.76 | 0.66 | **0.77** |
| | RP | PD-88, HC-88 | 0.69 | 0.70 | 0.74 | 0.62 | 0.79 | 0.73 | 0.73 | 0.80 | 0.61 | 0.75 | 0.73 | 0.73 | 0.72 | 0.73 | 0.79 | 0.71 | 0.71 | 0.69 | 0.71 | 0.77 | 0.73 | 0.73 | 0.73 | 0.71 | 0.79 |
| | TDU | PD-88, HC-88 | 0.73 | 0.72 | 0.68 | 0.74 | 0.81 | 0.75 | 0.75 | 0.80 | 0.66 | 0.82 | 0.76 | 0.75 | 0.73 | 0.78 | 0.83 | 0.74 | 0.74 | 0.79 | 0.73 | 0.82 | 0.76 | **0.76** | 0.73 | 0.73 | **0.84** |
| CzechPD | SS | PD-20, HC-15 | 0.73 | 0.71 | 0.70 | 0.90 | 0.76 | 0.76 | 0.73 | 0.75 | 0.90 | **0.79** | 0.77 | 0.78 | 0.80 | 0.80 | 0.78 | 0.79 | 0.79 | 0.75 | 0.75 | 0.70 | 0.80 | **0.80** | 0.80 | 0.85 | 0.78 |
| | RP | PD-20, HC-15 | 0.80 | **0.77** | 0.75 | 0.70 | 0.80 | 0.78 | 0.71 | 0.75 | 0.90 | 0.90 | 0.71 | 0.71 | 0.70 | 0.65 | 0.85 | 0.71 | 0.71 | 0.70 | 0.70 | **0.85** | 0.73 | 0.72 | 0.75 | 0.70 | 0.85 |
| GITA | SS | PD-49, HC-50 | 0.51 | 0.50 | 0.40 | 0.60 | 0.50 | 0.54 | 0.54 | 0.65 | 0.64 | 0.57 | 0.55 | 0.51 | 0.55 | 0.52 | 0.57 | 0.55 | 0.55 | 0.53 | 0.56 | 0.57 | 0.56 | **0.56** | 0.55 | 0.56 | **0.60** |
| | RP | PD-49, HC-50 | 0.67 | 0.63 | 0.68 | 0.78 | 0.72 | 0.58 | 0.58 | 0.51 | 0.80 | **0.73** | 0.63 | 0.62 | 0.63 | 0.62 | 0.69 | 0.70 | **0.70** | 0.63 | 0.66 | 0.71 | 0.62 | 0.62 | 0.61 | 0.66 | 0.72 |
| | TDU | PD-50, HC-50 | 0.63 | 0.62 | 0.62 | 0.64 | 0.68 | 0.60 | 0.60 | 0.46 | 0.86 | 0.72 | 0.69 | 0.67 | 0.76 | 0.66 | 0.75 | 0.69 | 0.69 | 0.80 | 0.68 | **0.77** | 0.72 | **0.72** | 0.78 | 0.68 | 0.74 |
| Italian | RP | PD-24, HC-36 | 0.93 | 0.91 | 0.88 | 0.98 | 0.98 | 0.90 | 0.90 | 0.85 | 0.94 | 0.90 | 0.95 | **0.93** | 0.92 | 0.98 | 0.95 | 0.92 | 0.92 | 0.87 | 0.92 | **0.98** | 0.90 | 0.90 | 0.88 | 0.92 | 0.95 |
| | TDU | PD-22, HC-21 | 0.88 | 0.90 | 0.79 | 0.95 | 0.92 | 0.87 | 0.87 | 0.82 | 0.90 | 0.93 | 0.88 | 0.88 | 0.88 | 0.90 | 0.98 | 0.98 | **0.98** | 0.97 | 1.00 | **1.00** | 0.92 | 0.92 | 0.86 | 1.00 | 0.98 |

| **Multi-lingual** | | | | | | | | | | | | | | | | | | | | | | | | | | | |
|---|---|---|---|---|---|---|---|---|---|---|---|---|---|---|---|---|---|---|---|---|---|---|---|---|---|---|---|
| Test | Task | spk# | SVM | | | | | KNN | | | | | Random Forest | | | | | XGBoost | | | | | Bagging | | | | |
| | | | ACC | F1 | SPE | SEN | AUC | ACC | F1 | SPE | SEN | AUC | ACC | F1 | SPE | SEN | AUC | ACC | F1 | SPE | SEN | AUC | ACC | F1 | SPE | SEN | AUC |
| NLS | SS | PD-23, HC-27 | 0.80 | **0.82** | 0.79 | 0.71 | **0.83** | 0.73 | 0.71 | 0.70 | 0.85 | 0.80 | 0.64 | 0.67 | 0.72 | 0.57 | 0.77 | 0.68 | 0.70 | 0.62 | 0.69 | 0.77 | 0.68 | 0.70 | 0.71 | 0.57 | 0.76 |
| | RP | PD-23, HC-27 | 0.66 | 0.65 | 0.75 | 0.57 | 0.78 | 0.60 | 0.60 | 0.57 | 0.87 | 0.72 | 0.70 | 0.68 | 0.67 | 0.68 | 0.78 | 0.72 | **0.72** | 0.78 | 0.77 | 0.77 | 0.70 | 0.70 | 0.75 | 0.77 | **0.82** |
| Neurovoz | TDU | PD-43, HC-46 | 0.84 | **0.86** | 0.93 | 0.77 | **0.90** | 0.81 | 0.81 | 0.60 | 0.92 | 0.86 | 0.81 | 0.83 | 0.90 | 0.70 | 0.90 | 0.80 | 0.80 | 0.86 | 0.74 | 0.80 | 0.81 | 0.82 | 0.88 | 0.72 | 0.89 |
| | SS | PD-20, HC-21 | 0.76 | 0.78 | 0.80 | 0.62 | **0.92** | 0.68 | 0.68 | 0.70 | 0.72 | 0.82 | 0.84 | **0.85** | 0.95 | 0.72 | 0.88 | 0.74 | 0.74 | 0.80 | 0.62 | 0.86 | 0.74 | 0.74 | 0.85 | 0.62 | 0.91 |
| GermanPD | SS | PD-88, HC-88 | 0.71 | 0.69 | 0.65 | 0.72 | 0.75 | 0.66 | 0.66 | 0.53 | 0.76 | 0.71 | 0.68 | 0.68 | 0.67 | 0.68 | 0.74 | 0.71 | **0.71** | 0.71 | 0.69 | **0.75** | 0.70 | 0.70 | 0.67 | 0.69 | 0.74 |
| | RP | PD-88, HC-88 | 0.69 | 0.66 | 0.67 | 0.78 | 0.79 | 0.66 | 0.66 | 0.66 | 0.76 | 0.77 | 0.73 | 0.71 | 0.72 | 0.77 | 0.81 | 0.76 | **0.76** | 0.70 | 0.71 | **0.83** | 0.73 | 0.73 | 0.71 | 0.71 | 0.81 |
| | TDU | PD-88, HC-88 | 0.73 | 0.71 | 0.70 | 0.75 | 0.76 | 0.73 | 0.73 | 0.64 | 0.79 | 0.79 | 0.76 | 0.74 | 0.72 | 0.81 | 0.81 | 0.73 | 0.73 | 0.66 | 0.76 | 0.80 | 0.76 | **0.76** | 0.67 | 0.76 | **0.80** |
| CzechPD | SS | PD-20, HC-15 | 0.72 | 0.78 | 0.85 | 0.45 | 0.72 | 0.68 | 0.68 | 0.75 | 0.65 | 0.80 | 0.76 | 0.79 | 0.85 | 0.50 | 0.72 | 0.82 | **0.82** | 0.90 | 0.50 | 0.65 | 0.76 | 0.76 | 0.80 | 0.50 | **0.85** |
| | RP | PD-20, HC-15 | 0.78 | **0.82** | 0.85 | 0.50 | 0.72 | 0.72 | 0.72 | 0.55 | 0.75 | 0.75 | 0.67 | 0.69 | 0.75 | 0.55 | **0.80** | 0.72 | 0.72 | 0.80 | 0.55 | 0.80 | 0.67 | 0.67 | 0.75 | 0.55 | 0.72 |
| GITA | SS | PD-49, HC-50 | 0.52 | 0.51 | 0.44 | 0.60 | 0.56 | 0.57 | 0.57 | 0.43 | 0.72 | 0.61 | 0.58 | 0.53 | 0.48 | 0.66 | 0.60 | 0.60 | **0.60** | 0.52 | 0.58 | **0.62** | 0.59 | 0.59 | 0.48 | 0.58 | 0.62 |
| | RP | PD-49, HC-50 | 0.58 | 0.53 | 0.61 | 0.68 | 0.69 | 0.62 | 0.62 | 0.40 | 0.84 | 0.72 | 0.66 | 0.63 | 0.63 | 0.66 | **0.75** | 0.66 | 0.66 | 0.59 | 0.68 | 0.74 | 0.67 | **0.67** | 0.61 | 0.68 | 0.71 |
| | TDU | PD-50, HC-50 | 0.66 | 0.62 | 0.54 | 0.70 | 0.73 | 0.60 | 0.60 | 0.36 | 0.88 | 0.71 | 0.68 | 0.66 | 0.66 | 0.72 | 0.74 | 0.69 | 0.69 | 0.62 | 0.74 | 0.77 | 0.71 | **0.71** | 0.66 | 0.74 | **0.78** |
| ItalianPVS | RP | PD-24, HC-36 | 0.85 | **0.86** | 0.87 | 0.79 | **0.93** | 0.72 | 0.72 | 0.57 | 0.84 | 0.85 | 0.77 | 0.74 | 0.80 | 0.76 | 0.86 | 0.85 | 0.85 | 0.80 | 0.79 | 0.88 | 0.82 | 0.82 | 0.80 | 0.79 | 0.88 |
| | TDU | PD-22, HC-21 | 0.84 | **0.87** | 0.90 | 0.75 | 0.86 | 0.71 | 0.71 | 0.39 | 0.87 | 0.80 | 0.81 | 0.85 | 0.88 | 0.67 | 0.80 | 0.86 | 0.86 | 0.88 | 0.67 | **0.87** | 0.79 | 0.79 | 0.93 | 0.67 | 0.76 |

| **Cross-lingual** | | | | | | | | | | | | | | | | | | | | | | | | | | | |
|---|---|---|---|---|---|---|---|---|---|---|---|---|---|---|---|---|---|---|---|---|---|---|---|---|---|---|---|
| Test | Task | spk# | SVM | | | | | KNN | | | | | Random Forest | | | | | XGBoost | | | | | Bagging | | | | |
| | | | ACC | F1 | SPE | SEN | AUC | ACC | F1 | SPE | SEN | AUC | ACC | F1 | SPE | SEN | AUC | ACC | F1 | SPE | SEN | AUC | ACC | F1 | SPE | SEN | AUC |
| NLS | SS | PD-23, HC-27 | 0.67 | 0.68 | 0.73 | 0.61 | 0.70 | 0.65 | 0.65 | 0.38 | 0.89 | 0.71 | 0.76 | 0.70 | 0.58 | 0.93 | 0.74 | 0.72 | 0.72 | 0.65 | 0.79 | 0.75 | 0.76 | **0.76** | 0.65 | 0.79 | **0.76** |
| | RP | PD-23, HC-27 | 0.56 | 0.67 | 0.96 | 0.22 | 0.69 | 0.54 | 0.54 | 0.83 | 0.30 | 0.62 | 0.66 | 0.71 | 0.91 | 0.44 | 0.79 | 0.72 | **0.72** | 0.87 | 0.59 | **0.79** | 0.64 | 0.64 | 0.87 | 0.59 | 0.79 |
| Neurovoz | TDU | PD-43, HC-46 | 0.66 | **0.72** | 0.86 | 0.43 | 0.72 | 0.70 | 0.70 | 0.84 | 0.72 | **0.76** | 0.57 | 0.69 | 0.96 | 0.04 | 0.71 | 0.60 | 0.69 | 0.94 | 0.39 | 0.71 | 0.56 | 0.56 | 0.95 | 0.39 | 0.67 |
| | SS | PD-20, HC-21 | 0.51 | 0.60 | 0.75 | 0.29 | 0.78 | 0.59 | 0.59 | 0.70 | 0.48 | 0.66 | 0.68 | **0.73** | 0.90 | 0.48 | **0.85** | 0.68 | 0.68 | 0.90 | 0.48 | 0.79 | 0.68 | 0.68 | 0.90 | 0.48 | 0.81 |
| GermanPD | SS | PD-88, HC-88 | 0.45 | 0.21 | 0.15 | 0.75 | 0.52 | 0.48 | 0.48 | 0.03 | 0.93 | 0.55 | 0.51 | 0.25 | 0.16 | 0.86 | 0.58 | 0.52 | 0.52 | 0.16 | 0.89 | 0.60 | 0.52 | **0.52** | 0.12 | 0.89 | **0.60** |
| | RP | PD-88, HC-88 | 0.48 | 0.26 | 0.70 | 0.78 | 0.52 | 0.52 | 0.52 | 0.67 | 0.99 | 0.54 | 0.48 | 0.22 | 0.65 | 0.82 | 0.51 | 0.53 | **0.53** | 0.65 | 0.82 | **0.60** | 0.51 | 0.51 | 0.68 | 0.82 | 0.52 |
| | TDU | PD-88, HC-88 | 0.53 | 0.66 | 0.92 | 0.11 | 0.52 | 0.52 | 0.52 | 0.30 | 0.82 | 0.63 | 0.57 | **0.66** | 0.83 | 0.38 | 0.63 | 0.58 | 0.58 | 0.77 | 0.38 | 0.62 | 0.56 | 0.56 | 0.67 | 0.38 | **0.64** |
| CzechPD | SS | PD-20, HC-15 | 0.60 | **0.67** | 0.70 | 0.86 | 0.68 | 0.43 | 0.43 | 0.20 | 0.81 | 0.64 | 0.43 | 0.50 | 0.40 | 0.82 | 0.68 | 0.49 | 0.49 | 0.10 | 0.77 | 0.69 | 0.43 | 0.43 | 0.20 | 0.77 | **0.73** |
| | RP | PD-20, HC-15 | 0.54 | 0.54 | 0.54 | 0.54 | 0.69 | 0.46 | 0.46 | 0.20 | 0.80 | 0.57 | 0.60 | 0.46 | 0.30 | 1.00 | 0.76 | 0.70 | **0.70** | 0.45 | 1.00 | **0.79** | 0.63 | 0.63 | 0.35 | 1.00 | 0.79 |
| GITA | SS | PD-49, HC-50 | 0.56 | 0.56 | 0.56 | 0.56 | 0.56 | 0.56 | 0.56 | 0.80 | 0.32 | 0.52 | 0.52 | **0.64** | 0.86 | 0.18 | **0.59** | 0.51 | 0.51 | 0.90 | 0.20 | 0.55 | 0.53 | 0.53 | 0.86 | 0.20 | 0.58 |
| | RP | PD-49, HC-50 | 0.48 | 0.65 | 1.00 | 0.00 | 0.42 | 0.49 | 0.49 | 0.94 | 0.04 | 0.45 | 0.49 | **0.66** | 0.98 | 0.02 | 0.43 | 0.51 | 0.51 | 1.00 | 0.04 | 0.50 | 0.52 | 0.52 | 0.98 | 0.04 | **0.55** |
| | TDU | PD-50, HC-50 | 0.53 | **0.65** | 0.67 | 0.20 | 0.64 | 0.63 | 0.63 | 0.52 | 0.52 | 0.65 | 0.53 | 0.32 | 0.29 | 0.84 | 0.62 | 0.53 | 0.53 | 0.26 | 0.88 | 0.62 | 0.54 | 0.54 | 0.26 | 0.88 | **0.66** |
| ItalianPVS | RP | PD-24, HC-36 | 0.63 | 0.66 | 0.79 | 0.67 | 0.78 | 0.65 | 0.65 | 0.83 | 0.58 | 0.75 | 0.72 | 0.54 | 0.50 | 0.92 | 0.71 | 0.77 | **0.77** | 0.46 | 0.94 | **0.76** | 0.75 | 0.75 | 0.46 | 0.94 | 0.74 |
| | TDU | PD-22, HC-21 | 0.70 | **0.74** | 0.77 | 0.94 | 0.74 | 0.64 | 0.64 | 0.58 | 0.94 | 0.77 | 0.68 | 0.65 | 0.58 | 0.93 | **0.84** | 0.64 | 0.64 | 0.46 | 0.96 | 0.83 | 0.68 | 0.68 | 0.54 | 0.96 | 0.83 |

Table 2: Results for mono-lingual, multi-lingual, and cross-lingual experiments using interpretable features. Results are reported for SS, RP and TDU tasks respectively. Accuracy (ACC), F-1 score, sensitivity (SEN), specificity (SPE), and area under the ROC (AUC) are reported. The best F1 and AUC across each row are emphasized in bold. For multi-lingual and cross-lingual experiments, shaded F1 and AUC represent an improvement $\geq 5\%$ with respect to mono-lingual experiments.

Table 3: Results for mono-lingual, multi-lingual, and cross-lingual experiments using x-vectors and TRILLsson features respectively. Results are reported for SS, RP and TDU tasks respectively. Accuracy (ACC), F1-score, sensitivity (SEN), specificity (SPE), and area under the ROC (AUC) are reported. The best F1 and AUC across each row are emphasized in bold. For multi-lingual and cross-lingual experiments, shaded F1 and AUC represent an improvement ≥5% with respect to mono-lingual experiments.

| **Mono-lingual** | | | | | | | | | | | | |
|---|---|---|---|---|---|---|---|---|---|---|---|---|
| Test | Task | spk# | x-vector | | | | | TRILLsson | | | | |
| | | | ACC | F1 | SPE | SEN | AUC | ACC | F1 | SPE | SEN | AUC |
| NLS | SS | PD-23, HC-27 | 0.65 | 0.65 | 0.55 | 0.77 | 0.74 | 0.78 | **0.78** | 0.76 | 0.81 | **0.82** |
| | RP | PD-23, HC-27 | 0.63 | 0.63 | 0.57 | 0.70 | 0.64 | 0.67 | **0.66** | 0.54 | 0.83 | **0.73** |
| Neurovoz | TDU | PD-43, HC-46 | 0.82 | 0.82 | 0.83 | 0.81 | 0.92 | 0.90 | **0.90** | 0.87 | 0.93 | **0.95** |
| | SS | PD-20, HC-21 | 0.76 | **0.75** | 0.86 | 0.65 | **0.75** | 0.73 | 0.73 | 0.71 | 0.75 | 0.74 |
| GermanPD | SS | PD-88, HC-88 | 0.74 | 0.74 | 0.76 | 0.73 | 0.81 | 0.80 | **0.80** | 0.78 | 0.81 | **0.84** |
| | RP | PD-88, HC-88 | 0.70 | 0.70 | 0.72 | 0.69 | 0.78 | 0.79 | **0.79** | 0.83 | 0.75 | **0.83** |
| | TDU | PD-88, HC-88 | 0.68 | 0.68 | 0.69 | 0.66 | 0.73 | 0.78 | **0.78** | 0.76 | 0.81 | **0.84** |
| CzechPD | SS | PD-20, HC-15 | 0.83 | **0.83** | 0.87 | 0.80 | 0.86 | 0.80 | 0.80 | 0.93 | 0.70 | **0.92** |
| | RP | PD-20, HC-15 | 0.77 | **0.77** | 0.80 | 0.75 | 0.83 | 0.77 | **0.77** | 0.87 | 0.70 | **0.85** |
| GITA | SS | PD-49, HC-50 | 0.76 | **0.76** | 0.76 | 0.76 | 0.83 | 0.76 | **0.76** | 0.78 | 0.73 | **0.84** |
| | RP | PD-49, HC-50 | 0.76 | 0.76 | 0.82 | 0.69 | 0.84 | 0.84 | **0.84** | 0.84 | 0.84 | **0.87** |
| | TDU | PD-50, HC-50 | 0.74 | 0.74 | 0.80 | 0.68 | 0.86 | 0.78 | **0.78** | 0.78 | 0.78 | **0.87** |
| ItalianPVS | RP | PD-24, HC-36 | 0.83 | 0.83 | 0.75 | 0.96 | **0.98** | 0.85 | **0.85** | 0.78 | 0.96 | 0.95 |
| | TDU | PD-22, HC-21 | 0.85 | 0.85 | 1.00 | 0.73 | **1.00** | 0.89 | **0.89** | 1.00 | 0.81 | 0.99 |
| **Multi-lingual** | | | | | | | | | | | | |
| Test | Task | spk# | x-vector | | | | | TRILLsson | | | | |
| | | | ACC | F1 | SPE | SEN | AUC | ACC | F1 | SPE | SEN | AUC |
| NLS | SS | PD-23, HC-27 | 0.78 | 0.78 | 0.76 | 0.81 | 0.84 | 0.85 | **0.85** | 0.79 | 0.92 | **0.92** |
| | RP | PD-23, HC-27 | 0.65 | 0.64 | 0.68 | 0.61 | 0.68 | 0.67 | **0.67** | 0.64 | 0.70 | **0.77** |
| Neurovoz | TDU | PD-43, HC-46 | 0.84 | **0.84** | 0.87 | 0.81 | 0.89 | 0.84 | **0.84** | 0.83 | 0.86 | **0.93** |
| | SS | PD-20, HC-21 | 0.71 | 0.70 | 0.81 | 0.60 | 0.80 | 0.76 | **0.75** | 0.86 | 0.65 | **0.89** |
| GermanPD | SS | PD-88, HC-88 | 0.76 | 0.76 | 0.77 | 0.75 | 0.81 | 0.77 | **0.77** | 0.81 | 0.74 | **0.84** |
| | RP | PD-88, HC-88 | 0.70 | 0.70 | 0.64 | 0.76 | 0.79 | 0.85 | **0.85** | 0.80 | 0.91 | **0.88** |
| | TDU | PD-88, HC-88 | 0.74 | 0.74 | 0.78 | 0.70 | 0.79 | 0.79 | **0.79** | 0.80 | 0.78 | **0.84** |
| CzechPD | SS | PD-20, HC-15 | 0.71 | 0.71 | 0.67 | 0.75 | 0.84 | 0.89 | **0.88** | 0.80 | 0.95 | **0.93** |
| | RP | PD-20, HC-15 | 0.69 | 0.68 | 0.60 | 0.75 | 0.81 | 0.91 | **0.91** | 0.87 | 0.95 | **0.90** |
| GITA | SS | PD-49, HC-50 | 0.76 | 0.76 | 0.80 | 0.71 | 0.80 | 0.81 | **0.81** | 0.80 | 0.82 | **0.91** |
| | RP | PD-49, HC-50 | 0.72 | 0.72 | 0.80 | 0.63 | 0.78 | 0.80 | **0.80** | 0.80 | 0.80 | **0.88** |
| | TDU | PD-50, HC-50 | 0.74 | 0.74 | 0.78 | 0.70 | 0.82 | 0.78 | **0.78** | 0.80 | 0.76 | **0.86** |
| ItalianPVS | RP | PD-24, HC-36 | 0.90 | **0.90** | 0.89 | 0.92 | **0.97** | 0.90 | **0.90** | 0.89 | 0.92 | **0.97** |
| | TDU | PD-22, HC-21 | 0.81 | 0.80 | 0.67 | 0.92 | 0.94 | 0.91 | **0.91** | 0.86 | 0.96 | **0.97** |
| **Cross-lingual** | | | | | | | | | | | | |
| Test | Task | spk# | x-vector | | | | | TRILLsson | | | | |
| | | | ACC | F1 | SPE | SEN | AUC | ACC | F1 | SPE | SEN | AUC |
| NLS | SS | PD-23, HC-27 | 0.69 | **0.68** | 0.52 | 0.88 | 0.77 | 0.58 | 0.53 | 0.24 | 0.96 | **0.89** |
| | RP | PD-23, HC-27 | 0.55 | **0.54** | 0.39 | 0.74 | 0.66 | 0.43 | 0.30 | 0.00 | 0.96 | **0.76** |
| Neurovoz | TDU | PD-43, HC-46 | 0.62 | 0.60 | 0.37 | 0.89 | 0.78 | 0.72 | **0.69** | 0.98 | 0.44 | **0.88** |
| | SS | PD-20, HC-21 | 0.71 | **0.71** | 0.67 | 0.75 | 0.77 | 0.68 | 0.65 | 0.95 | 0.40 | **0.90** |
| GermanPD | SS | PD-88, HC-88 | 0.68 | 0.68 | 0.70 | 0.65 | 0.74 | 0.75 | **0.74** | 0.60 | 0.90 | **0.84** |
| | RP | PD-88, HC-88 | 0.62 | **0.60** | 0.40 | 0.84 | 0.70 | 0.61 | 0.57 | 0.93 | 0.30 | **0.81** |
| | TDU | PD-88, HC-88 | 0.53 | **0.41** | 0.08 | 0.99 | 0.72 | 0.52 | 0.38 | 0.05 | 1.00 | **0.79** |
| CzechPD | SS | PD-20, HC-15 | 0.66 | 0.65 | 0.87 | 0.50 | 0.76 | 0.89 | **0.89** | 1.00 | 0.80 | **0.97** |
| | RP | PD-20, HC-15 | 0.63 | 0.62 | 0.93 | 0.40 | 0.74 | 0.74 | **0.74** | 0.93 | 0.60 | **0.90** |
| GITA | SS | PD-49, HC-50 | 0.56 | 0.55 | 0.46 | 0.65 | 0.66 | 0.82 | **0.82** | 0.92 | 0.71 | **0.89** |
| | RP | PD-49, HC-50 | 0.63 | **0.61** | 0.42 | 0.84 | 0.77 | 0.61 | 0.55 | 0.24 | 0.98 | **0.80** |
| | TDU | PD-50, HC-50 | 0.62 | 0.62 | 0.56 | 0.68 | 0.70 | 0.70 | **0.69** | 0.50 | 0.90 | **0.82** |
| ItalianPVS | RP | PD-24, HC-36 | 0.80 | **0.79** | 0.86 | 0.71 | 0.88 | 0.75 | 0.69 | 1.00 | 0.38 | **0.94** |
| | TDU | PD-22, HC-21 | 0.64 | 0.64 | 0.67 | 0.62 | 0.68 | 0.66 | **0.66** | 0.86 | 0.50 | **0.83** |

Table 4: Results for mono-lingual, multi-lingual, and cross-lingual experiments using Wav2Vec 2.0. and HuBERT features, respectively. Results are reported for SS, RP, and TDU tasks respectively. Accuracy (ACC), F-1 score, sensitivity (SEN), specificity (SPE), and area under the ROC (AUC) are reported. The best F1 and AUC across each row are emphasized in bold. For multi-lingual and cross-lingual experiments, shaded F1 and AUC represent an improvement ≥5% with respect to mono-lingual experiments. For Wav2Vec 2.0., we report results obtained using layer 4, and for Hubert, those obtained using layer 7. We did so as we achieved the highest performances using representations extracted from these layers.

| **Mono-lingual** | | | | | | | | | | | | |
|---|---|---|---|---|---|---|---|---|---|---|---|---|
| Test | Task | spk# | Wav2Vec 2.0. | | | | | HuBERT | | | | |
| | | | ACC | F1 | SPE | SEN | AUC | ACC | F1 | SPE | SEN | AUC |
| NLS | SS | PD-23, HC-27 | 0.78 | **0.78** | 0.83 | 0.73 | **0.77** | 0.67 | 0.62 | 0.76 | 0.58 | 0.70 |
| | RP | PD-23, HC-27 | 0.63 | **0.63** | 0.64 | 0.61 | 0.60 | 0.63 | 0.60 | 0.64 | 0.61 | **0.62** |
| Neurovoz | TDU | PD-43, HC-46 | 0.91 | **0.91** | 0.89 | 0.93 | **0.96** | 0.90 | 0.90 | 0.89 | 0.91 | **0.96** |
| | SS | PD-20, HC-21 | 0.66 | 0.66 | 0.62 | 0.70 | 0.67 | 0.73 | **0.74** | 0.67 | 0.80 | **0.78** |
| GermanPD | SS | PD-88, HC-88 | 0.76 | 0.76 | 0.77 | 0.74 | 0.85 | 0.78 | **0.78** | 0.81 | 0.76 | **0.86** |
| | RP | PD-88, HC-88 | 0.78 | **0.78** | 0.78 | 0.78 | 0.83 | 0.78 | 0.77 | 0.83 | 0.74 | **0.88** |
| | TDU | PD-88, HC-88 | 0.75 | 0.75 | 0.75 | 0.75 | 0.79 | 0.79 | **0.78** | 0.85 | 0.72 | **0.88** |
| CzechPD | SS | PD-20, HC-15 | 0.86 | **0.86** | 0.93 | 0.80 | 0.88 | 0.80 | 0.81 | 0.87 | 0.75 | **0.91** |
| | RP | PD-20, HC-15 | 0.83 | 0.83 | 0.87 | 0.80 | **0.89** | 0.83 | **0.84** | 0.87 | 0.80 | 0.83 |
| GITA | SS | PD-49, HC-50 | 0.86 | **0.86** | 0.90 | 0.82 | **0.90** | 0.83 | 0.82 | 0.86 | 0.79 | 0.89 |
| | RP | PD-49, HC-50 | 0.76 | **0.76** | 0.76 | 0.76 | **0.86** | 0.75 | 0.74 | 0.76 | 0.74 | 0.79 |
| | TDU | PD-50, HC-50 | 0.82 | **0.82** | 0.82 | 0.82 | **0.89** | 0.79 | 0.78 | 0.82 | 0.76 | 0.85 |
| ItalianPVS | RP | PD-24, HC-36 | 0.83 | 0.83 | 0.75 | 0.96 | 0.96 | 0.85 | **0.84** | 0.78 | 0.96 | **0.97** |
| | TDU | PD-22, HC-21 | 0.87 | **0.87** | 1.00 | 0.77 | **1.00** | 0.85 | 0.84 | 1.00 | 0.73 | **1.00** |
| **Multi-lingual** | | | | | | | | | | | | |
| Test | Task | spk# | Wav2Vec 2.0. | | | | | HuBERT | | | | |
| | | | ACC | F1 | SPE | SEN | AUC | ACC | F1 | SPE | SEN | AUC |
| NLS | SS | PD-23, HC-27 | 0.80 | **0.80** | 0.79 | 0.81 | **0.80** | 0.65 | 0.64 | 0.66 | 0.65 | 0.72 |
| | RP | PD-23, HC-27 | 0.76 | **0.76** | 0.86 | 0.65 | **0.72** | 0.67 | 0.62 | 0.71 | 0.60 | 0.71 |
| Neurovoz | TDU | PD-43, HC-46 | 0.91 | **0.91** | 0.89 | 0.93 | **0.96** | 0.84 | 0.84 | 0.83 | 0.86 | 0.93 |
| | SS | PD-20, HC-21 | 0.76 | 0.75 | 0.86 | 0.65 | **0.84** | 0.78 | **0.77** | 0.81 | 0.75 | 0.83 |
| GermanPD | SS | PD-88, HC-88 | 0.82 | **0.82** | 0.75 | 0.89 | **0.86** | 0.77 | 0.77 | 0.79 | 0.75 | **0.86** |
| | RP | PD-88, HC-88 | 0.77 | 0.77 | 0.73 | 0.82 | **0.86** | 0.80 | **0.79** | 0.81 | 0.78 | 0.85 |
| | TDU | PD-88, HC-88 | 0.76 | 0.76 | 0.76 | 0.75 | **0.82** | 0.80 | **0.80** | 0.81 | 0.80 | 0.79 |
| CzechPD | SS | PD-20, HC-15 | 0.83 | 0.83 | 0.80 | 0.85 | **0.92** | 0.83 | **0.86** | 0.73 | 0.90 | 0.87 |
| | RP | PD-20, HC-15 | 0.83 | 0.82 | 0.73 | 0.90 | **0.93** | 0.86 | **0.87** | 0.87 | 0.85 | 0.92 |
| GITA | SS | PD-49, HC-50 | 0.76 | 0.76 | 0.78 | 0.73 | **0.85** | 0.79 | **0.77** | 0.84 | 0.73 | 0.83 |
| | RP | PD-49, HC-50 | 0.82 | **0.82** | 0.84 | 0.80 | **0.88** | 0.73 | 0.73 | 0.72 | 0.73 | 0.81 |
| | TDU | PD-50, HC-50 | 0.75 | 0.75 | 0.82 | 0.68 | **0.86** | 0.79 | **0.78** | 0.84 | 0.74 | 0.85 |
| ItalianPVS | RP | PD-24, HC-36 | 0.90 | 0.90 | 0.86 | 0.96 | **0.98** | 0.93 | **0.92** | 0.92 | 0.96 | **0.98** |
| | TDU | PD-22, HC-21 | 0.91 | 0.91 | 0.81 | 1.00 | **0.99** | 0.94 | **0.95** | 0.86 | 1.00 | **0.99** |
| **Cross-lingual** | | | | | | | | | | | | |
| Test | Task | spk# | Wav2Vec 2.0. | | | | | HuBERT | | | | |
| | | | ACC | F1 | SPE | SEN | AUC | ACC | F1 | SPE | SEN | AUC |
| NLS | SS | PD-23, HC-27 | 0.73 | **0.72** | 0.83 | 0.62 | **0.76** | 0.65 | 0.71 | 0.45 | 0.88 | 0.70 |
| | RP | PD-23, HC-27 | 0.63 | 0.61 | 0.75 | 0.48 | 0.67 | 0.67 | **0.64** | 0.68 | 0.65 | **0.74** |
| Neurovoz | TDU | PD-43, HC-46 | 0.73 | 0.72 | 0.57 | 0.91 | **0.89** | 0.78 | **0.77** | 0.76 | 0.79 | 0.87 |
| | SS | PD-20, HC-21 | 0.63 | 0.60 | 0.33 | 0.95 | 0.78 | 0.71 | **0.62** | 0.90 | 0.50 | **0.81** |
| GermanPD | SS | PD-88, HC-88 | 0.72 | **0.71** | 0.81 | 0.63 | 0.78 | 0.68 | 0.61 | 0.86 | 0.50 | **0.79** |
| | RP | PD-88, HC-88 | 0.65 | 0.62 | 0.36 | 0.94 | **0.78** | 0.69 | **0.67** | 0.74 | 0.64 | 0.75 |
| | TDU | PD-88, HC-88 | 0.62 | 0.57 | 0.27 | 0.97 | **0.78** | 0.58 | **0.70** | 0.19 | 0.96 | 0.76 |
| CzechPD | SS | PD-20, HC-15 | 0.89 | **0.89** | 1.00 | 0.80 | **0.92** | 0.74 | 0.76 | 0.80 | 0.70 | 0.85 |
| | RP | PD-20, HC-15 | 0.77 | 0.77 | 0.93 | 0.65 | **0.90** | 0.80 | **0.82** | 0.80 | 0.80 | 0.87 |
| GITA | SS | PD-49, HC-50 | 0.72 | **0.70** | 0.84 | 0.59 | **0.73** | 0.67 | 0.65 | 0.72 | 0.61 | 0.69 |
| | RP | PD-49, HC-50 | 0.69 | 0.69 | 0.64 | 0.73 | **0.79** | 0.70 | **0.71** | 0.66 | 0.73 | 0.75 |
| | TDU | PD-50, HC-50 | 0.61 | 0.60 | 0.44 | 0.78 | 0.74 | 0.66 | **0.66** | 0.66 | 0.66 | **0.78** |
| ItalianPVS | RP | PD-24, HC-36 | 0.75 | **0.74** | 0.81 | 0.67 | 0.82 | 0.80 | 0.68 | 0.97 | 0.54 | **0.91** |
| | TDU | PD-22, HC-21 | 0.64 | **0.63** | 0.57 | 0.69 | **0.70** | 0.62 | 0.55 | 0.86 | 0.42 | **0.70** |
